# Risk of *Plasmodium vivax* recurrences follows a 30-70 rule and indicates relapse heterogeneity in the population

**DOI:** 10.1101/2022.05.18.22275180

**Authors:** Eva Stadler, Deborah Cromer, Somya Mehra, Adeshina I Adekunle, Jennifer A Flegg, Nicholas M Anstey, James A Watson, Cindy S Chu, Ivo Mueller, Leanne J Robinson, Timothy E Schlub, Miles P Davenport, David S Khoury

## Abstract

A key characteristic of *Plasmodium vivax* parasites is their ability to adopt a latent liver-stage form called hypnozoites, able to cause relapse of infection months or years after a primary infection. Relapses of infection through hypnozoite activation are a major contributor to blood-stage infections in *P vivax* endemic regions and are thought to be influenced by factors such as febrile infections, immunity, and transmission intensity. Some of these factors may cause temporary changes in hypnozoite activation over time, leading to ‘temporal heterogeneity’ in reactivation risk. In addition, variation in exposure to infection may be a longer-term characteristic of individuals that leads to ‘population heterogeneity’ in hypnozoite activation. We analyze data on risk of *P vivax* in two previously published data sets from Papua New Guinea and the Thailand-Myanmar border region. Modeling different mechanisms of reactivation risk, we find strong evidence for population heterogeneity, with 30% of patients having almost 70% of all *P vivax* infections. Model fitting and data analysis indicates that individual variation in relapse risk is a primary source of heterogeneity of *P vivax* risk of recurrences.

## Introduction

*Plasmodium vivax* is a major cause of clinical malaria with about 4.5 million cases in 2020 [1]. An important feature of *P vivax* malaria is the ability of the parasite to form latent liver-stage parasites (hypnozoites), which can later activate and initiate blood stage infection in the absence of new mosquito inoculation [2-6]. It has been estimated that 66% to 96% of *P vivax* blood-stage infections are relapses caused by the activation of hypnozoites [2, 4, 7-9]. These and other studies have highlighted that hypnozoite reactivation is a major source of observed blood-stage infections and presents a major barrier to effective control and eradication of *P vivax* malaria [4, 5, 7, 9]. Although the drug primaquine can effectively clear hypnozoites, its use has been limited in part because it can induce severe haemolysis in glucose-6-phosphate dehydrogenase (G6PD) deficient individuals [2, 4, 5, 9].

Several factors are thought to influence the timing of hypnozoite activation leading to relapse. In particular, recent infection with *P falciparum* or another infectious agent, and other factors may cause a temporary elevation in the risk of relapse [8, 10, 11]. Besides these temporal factors influencing the risk of relapse (‘temporal heterogeneity’), other factors such as transmission intensity, variation in latency due to differences in *P vivax* strains (e.g. temperate and tropical latency phenotypes) [6, 12], host immunity and age are also known to influence the pattern of blood-stage infections [4, 6, 10, 13, 14]. Further, across a patient population, individuals are likely to harbor different numbers of hypnozoites due to differences in infection risk and a skewed distribution of sporozoite numbers inoculated [6, 15, 16], use of primaquine for radical cure and CYP2D6 polymorphisms causing treatment failures in some individuals [17], or variation in infection susceptibility [18] (which for example may be due to G6PD deficiency [19]). Given these differences, we might expect that individuals within a population may vary significantly in the number of hypnozoites they harbor, which is expected to contribute directly to their risk of hypnozoite reactivation and *P vivax* relapse. In particular, studies of *P cynomolgi* in Rhesus macaques have shown that there is a correlation between the sporozoite inoculation and relapse frequency [20]. Thus, more exposed individuals within a community are expected to be more likely to have a larger hypnozoite reservoir and to have more frequent relapses [6, 16]. Indeed, heterogeneity in malaria transmission, infections, and *P vivax* recurrences (i.e., *P vivax* infections that are either due to a new, mosquito-borne infection or a relapse) have been described previously [13, 14, 18]. For example, Chu et al. found that all recurrences observed after day 35 post enrolment occurred in only 12% of individuals [21]. This is consistent with studies of *P falciparum* infection, in which Cooper et al. found evidence that 20% of the population accounted for around 80% of transmission events [22].

Despite evidence for heterogeneity in *P vivax* relapses, much of the mathematical modelling of *P vivax* recurrences to date has assumed either constant or periodic recurrence rates [7, 8, 15, 23, 24], with some exceptions. In particular, Taylor et al. have included inter-individual heterogeneity in their time-to-event modelling by including a random effect that determines the probability of the next event being a relapse or a new infection [8]. Further, other models have included age (a proxy for immunity) as a source of heterogeneity [4, 14], and a recent study incorporated the concept of variable risk in the form of a ‘high susceptibility’ subpopulation that has a higher risk of recurrence [14]. Here, we seek direct evidence of temporal and population heterogeneity in *P vivax* recurrences in two previously published studies [4, 8, 21, 25]. The first study by Robinson et al. [4] contains data from a randomized placebo-controlled trial of blood-plus liver-stage drugs in Papua New Guinean children. The second data set consists of two studies by Chu et al. [21, 25] and was made available by Taylor et al. [8]. Chu et al. conducted randomised trials in patients from the Thailand-Myanmar border region with symptomatic *P vivax* malaria, treated them with various therapies, and recorded their *P vivax* recurrences over a one-year follow-up [21, 25]. A key difference in these studies is that the Robinson et al. study recruited individuals from communities regardless of whether they were infected, whereas the Chu et al. studies focus on treatment of individuals with symptomatic *P vivax* malaria at enrolment. These datasets provide an excellent opportunity for modelling of *P vivax* recurrences in areas with a short latency *P vivax* phenotype. We compare whether a model that considers (i) only a constant risk in relapse, (ii) temporarily increased relapse risk, (iii) individual variation in relapse risk, or (iv) both temporal and population heterogeneity, best explains the pattern of relapses observed in these trials. The evidence suggests that population heterogeneity in relapse risk is a primary determinant of relapse patterns. We speculate that this population heterogeneity in relapse is likely mediated by different hypnozoite burdens in individuals due to variation in individual exposure to inoculation with sporozoites. This agrees also with the observation that in the Thailand-Myanmar data, almost 70% of all *P vivax* infections occur in only 30% of the patients who are treated only for blood-stage infections [6, 26]. In addition to population heterogeneity, we find evidence that temporal heterogeneity may also contribute to the overall relapse kinetics in these studies, albeit to a lesser extent.

## Results

### Risk of relapse decreases with time since last event

To understand the pattern and timing of relapses, we analyzed and modeled two datasets, from Papua New Guinea (published by Robinson et al. [4]) and from the Thailand-Myanmar border region [21, 25] (published by Taylor et al. [8], see Methods and original publications for further details on the data). We compared the time to first *P vivax* infection or recurrence in individuals treated for blood-stage *P vivax* infection with or without primaquine treatment to eliminate hypnozoites (***Figure 1A, B***, see also ***Supplementary Table S9***). To estimate the relapse rate, we assumed that patients with primaquine treatment have only new infections but those treated only for blood-stage infection have both new infections and relapses (as in previous studies [7, 9]). The weekly incidence rates for patients who did not receive radical cure and thus had both relapses and new infections were analyzed. This suggests that the relapse rate was non-constant (***Figure 1C, D***). For example, in the Thailand-Myanmar data, 43.6% of the individuals at risk were observed to have an infection between day 30 and day 60, but only 26.6% of the individuals at risk at day 60 had an infection between day 60 and 90. These observations indicate that the relapse rate is non-constant over time but decreasing.

**Figure 1.**
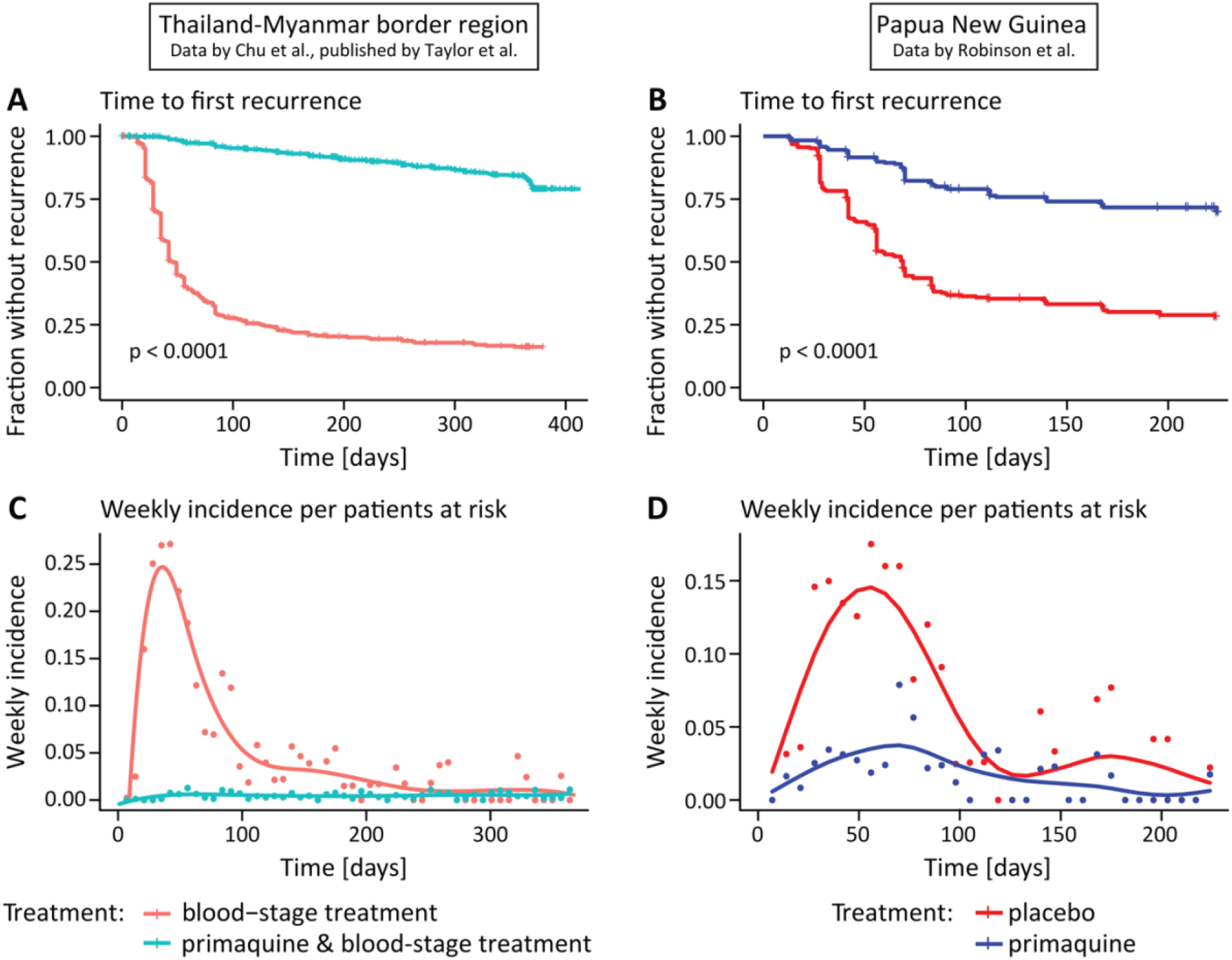
Time to first recurrence and weekly incidence per patients at risk in the Thailand-Myanmar and Papua New Guinea data. (**A, B**) Time from enrolment to the first recurrence for patients who received blood-stage treatment only (red) and patients who received primaquine and blood-stage treatment (blue). In the Papua New Guinea data, all patients (n = 504) received blood-stage treatment and either a placebo (red) or primaquine (blue). The time to first recurrence in the different treatment groups was compared using a log-rank test (p<0.0001 for both data sets). For the time to first recurrence in the Thailand-Myanmar data (n = 1298) by treatment and study see ***Supplementary Fig. S8***. (**C, D**) Weekly incidence per patients at risk for patients treated with primaquine and blood-stage treatment (blue dots) and blood-stage treatment only (red dots). The weekly incidence per patients at risk is the number of patients that had a recurrence within the current week divided by the number of patients who were at risk (i.e., the patients who have not yet had a recurrence) at the beginning of the week. The curves are fitted to the weekly incidence per patients at risk (using splines).

### Temporal and population heterogeneity in relapse risk as explanations of non-constant recurrence rates

We next consider models with different types of heterogeneity of relapse rates. First, we consider temporal heterogeneity, i.e., temporal variation of the relapse risk. Both the Thailand-Myanmar and the Papua New Guinea data show that the risk of a blood-stage infection decreases after an initial peak (***Figure 1***). This change in relapse risk may be due to, e.g., seasonal variations in the relapse risk [6, 12] or more rapid relapse after a recent infection [6, 10]. Therefore, we developed a model that allowed the relapse rate to decrease over time (see Materials and Methods). We compared this to a model with a constant rate of new infections and relapses. Both models allowed for a prophylactic period after treatment followed by new infections at a constant rate and either a constant relapse rate or a decreasing relapse rate (Materials and Methods). We found that the temporal heterogeneity model provided a significantly better fit to the data than the constant reactivation model (***Figure 2***, AIC differences of 190 and 44 in the Thailand-Myanmar and Papua New Guinea data, respectively, see ***Supplementary Fig. S2*** and ***Supplementary Fig. S10***).

**Figure 2.**
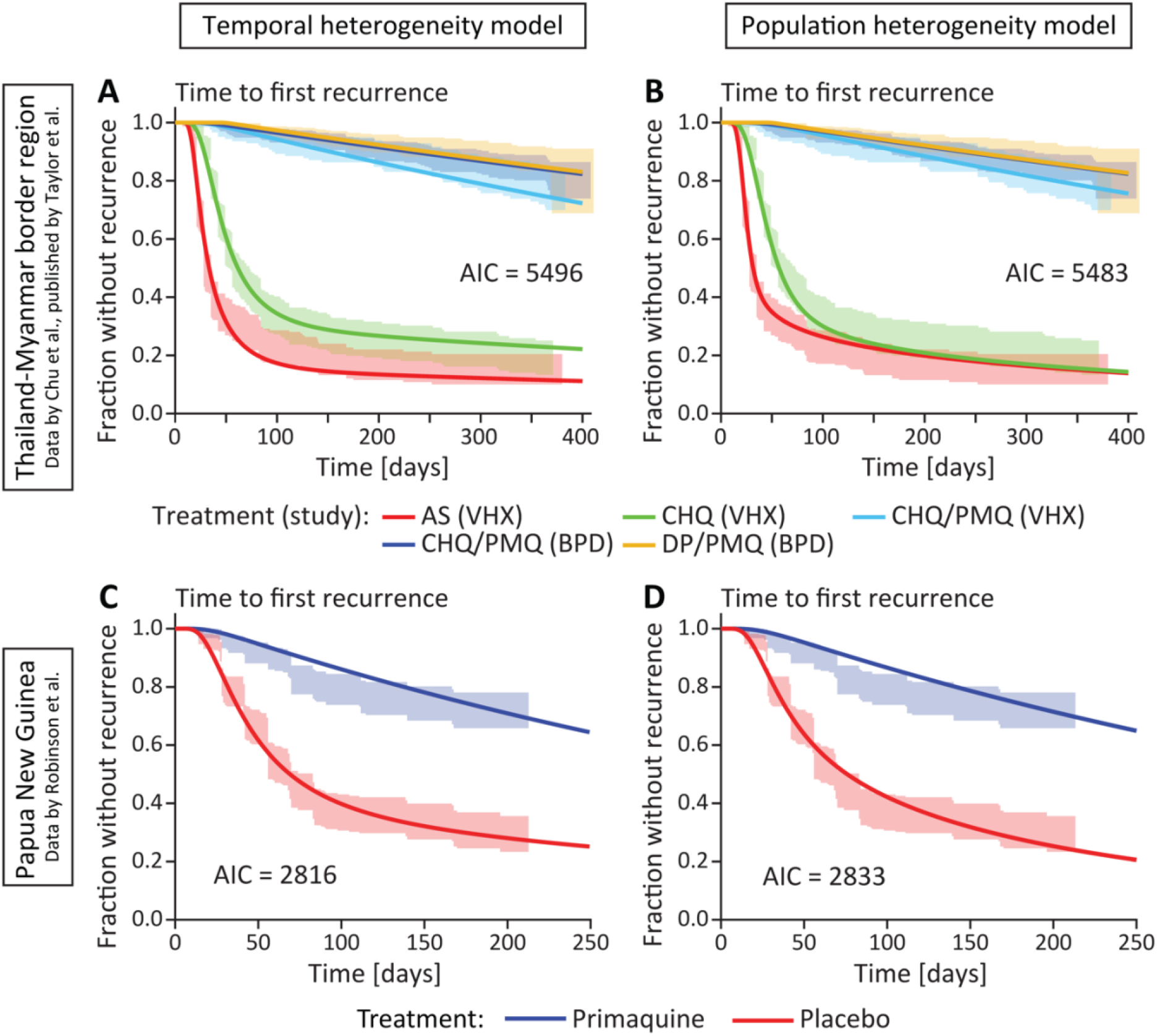
Fitting models of temporal and population heterogeneity to the data. The left column (**A** and **C**) shows the fit of the temporal heterogeneity model and the right column (**B** and **D**) shows the fit of the population heterogeneity model. The lines are the models fitted to the data and the shaded areas are the 95% confidence regions from the data. The models were fitted using a maximum likelihood approach (see Materials and Methods). (**A, B**) Fit of the heterogeneity models for each antimalarial treatment and study in the Thailand-Myanmar data. Abbreviations: AS artesunate, CHQ chloroquine, CHQ/PMQ chloroquine and primaquine, DP/PMQ dihydroartemisinin-piperaquine and primaquine, VHX Vivax History study, BPD best Primaquine Dose study. (**C, D**) Fit of the heterogeneity models to all Papua New Guinea data. For a fit to the Papua New Guinea data grouped by village see ***Supplementary Fig. S3*** and ***Supplementary Fig. S4***.

We also wanted to consider the impact of population heterogeneity in relapse rates, where individuals differ in their relapse rate due to factors such as their exposure, number of hypnozoites, age, or immunity. Importantly, this model is phenomenological in describing a distribution in relapse risk and does not attempt to explicitly model the mechanisms of population heterogeneity. The population heterogeneity model also provided a significantly better fit to the data than the constant relapse model (***Figure 2***, AIC differences of 203 and 27 in the Thailand Myanmar and Papua New Guinea data, respectively, see ***Supplementary Fig. S2*** and ***Supplementary Fig. S10***).

Comparing the temporal and population heterogeneity models, for both data sets, the two heterogeneity models both fit the data reasonably well with similar AIC differences. The model incorporating population heterogeneity provides a slightly better fit to the Thailand-Myanmar data (AIC difference of 13), and the temporal heterogeneity model provides a better fit to the Papua New Guinea data (AIC difference of 17). Thus, it is not clear whether temporal or population heterogeneity may be the more important source of heterogeneity in the ‘time-to-first-infection’ data (***Figure 2***).

### Simulating temporal and population heterogeneity

It is clear that simple fitting of time-to-first infection data cannot distinguish between temporal and population heterogeneity with the available data. To further explore the temporal and population heterogeneity interpretations of *P vivax* infection patterns, we developed a simulation of these processes based on the Thailand-Myanmar data model fits (see Methods and Supplementary methods). These simulations highlighted an important difference between these mechanisms in the time-to-second recurrence. That is, in the temporal heterogeneity model we observe that the time to first recurrence and time from first to second recurrence have a very weak negative correlation due to censoring after one year (***Table 1***). However, in the population heterogeneity model simulations, the time from enrollment to the first recurrence (time-to-first) and the time from the first to the second recurrence (time-to-second) in individuals are strongly positively correlated (***Table 1***). Thus, a key feature that differentiates these models is whether there is a correlation between time-to-first and time-to-second recurrence. To investigate this in the data, we fitted both models to time-to-first and time-to-second recurrence data and found that the population heterogeneity model provided a better fit of the data (***Figure 3***). To better understand this result, we also grouped individuals into quartiles based on the time to their first recurrence (***Figure 4B***), and plotted the time to their second recurrence for each group (***Figure 4C-D***). We found a clear correlation in the time between first and second recurrence (***Supplementary Fig. S7***, see also ***Supplementary Fig. S5, Table S11***, and ***Table S13***). This indicates that population heterogeneity in relapse risk is a major determinant of relapse patterns in the Thailand-Myanmar data. The observation of population heterogeneity in the risk of relapse would be consistent with individuals carrying variable numbers of hypnozoites and therefore experiencing different frequencies of relapse, which may occur because of random variation in inoculum size from infection or due to some individuals having higher exposure to primary *P vivax* infection.

**Table 1.**
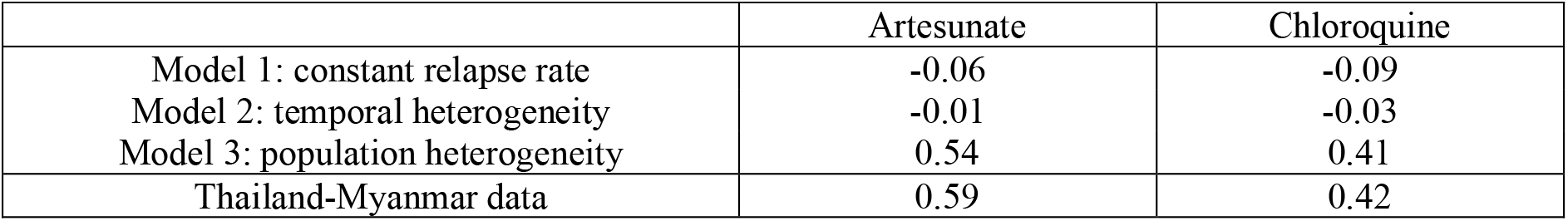
Spearman correlation between time to first recurrence and time from first to second recurrence in the simulated data and the Thailand-Myanmar data excluding censored data. All 1,000,000 simulated individuals who had at least two recurrences during the 1-year-simulation were used to compute the Spearman correlation. Note that the very weak negative correlation for models 1 and 2 is most likely due the censoring in the simulated data after 1 year of follow-up (see **Table S12** for details). All p-values are below 0.0001 due to the large number of simulations. For the Thailand-Myanmar data, we show here the Spearman correlation for artesunate or chloroquine treated individuals, excluding censored individuals (for the correlations of all individuals including censored data see **Table S11**).

|  | Artesunate | Chloroquine |
| --- | --- | --- |
| Model 1: constant relapse rate | -0.06 | -0.09 |
| Model 2: temporal heterogeneity | -0.01 | -0.03 |
| Model 3: population heterogeneity | 0.54 | 0.41 |
| Thailand-Myanmar data | 0.59 | 0.42 |

**Figure 3.**
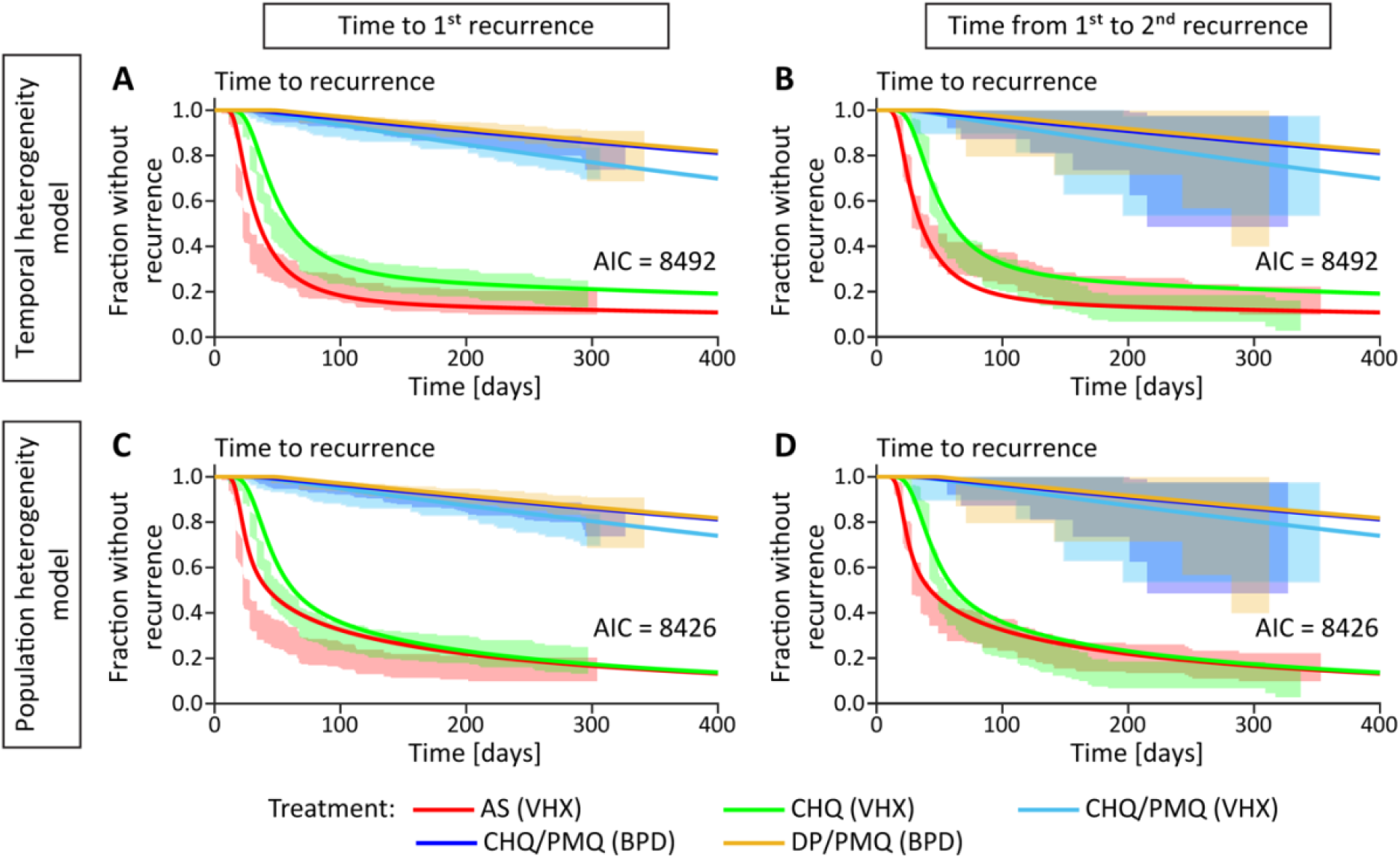
Fitting the temporal and population heterogeneity models to the first and second recurrence time in the Thailand-Myanmar data. Both recurrence times are fitted simultaneously (see Methods and Supplementary methods). The lines are the models fitted to the data and the shaded areas are the 95% confidence regions from the data. The left column shows the fit to the first recurrence time and the right column the fit to the time from the first to the second recurrence. The first row shows the temporal heterogeneity model fit and the second row the population heterogeneity model fit. Abbreviations: AS artesunate, CHQ chloroquine, CHQ/PMQ chloroquine and primaquine, DP/PMQ dihydroartemisinin-piperaquine and primaquine, VHX Vivax History study, BPD best Primaquine Dose study.

**Figure 4.**
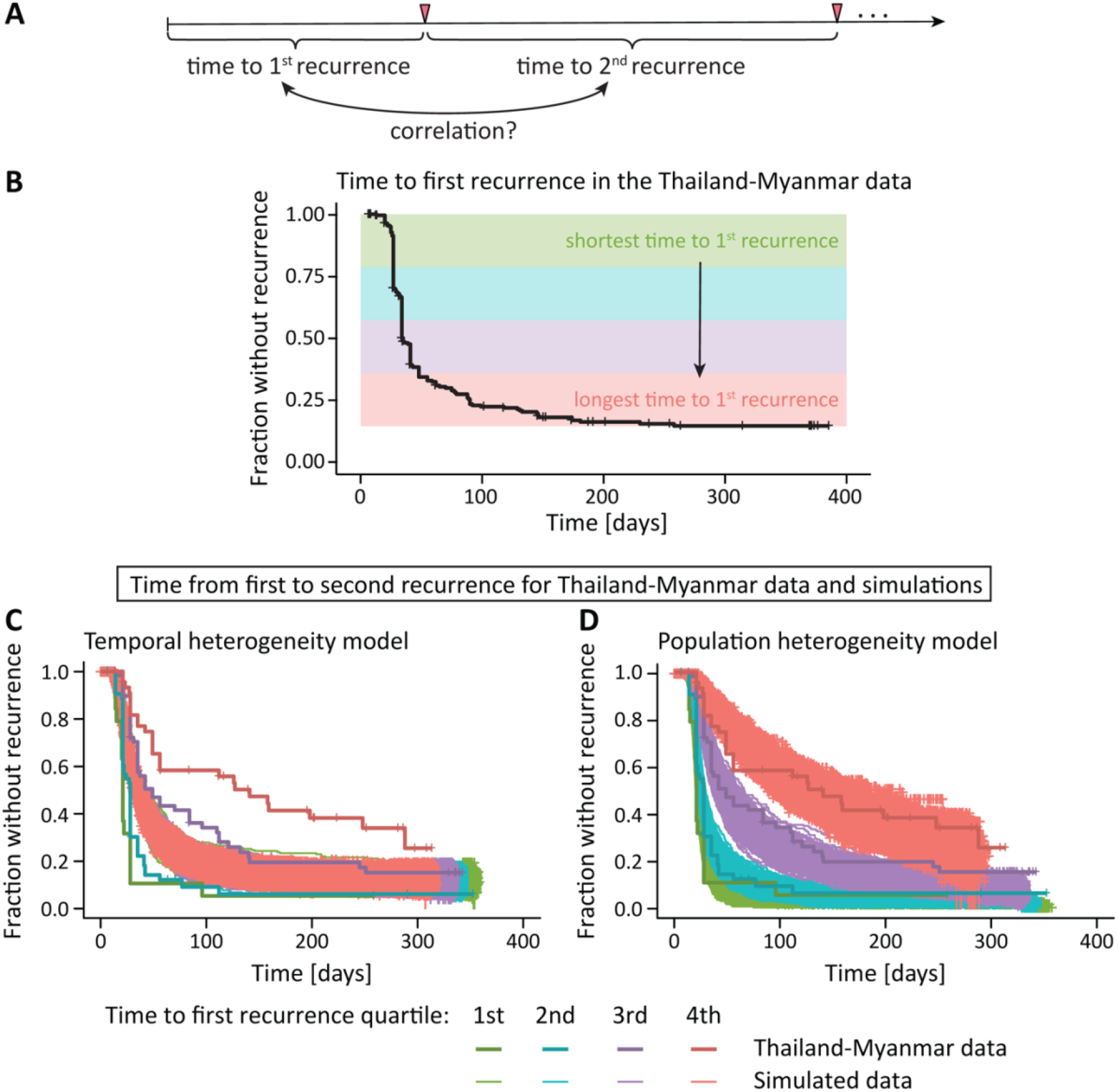
Association between different recurrence times. (**A**) Since we have multiple within patient recurrence times in the Thailand-Myanmar data, we can estimate the correlation between the time to first recurrence and the time to the second recurrence. (**B**) All individuals in the Thailand-Myanmar data who were treated with artesunate are grouped by their time to first recurrence quartiles, from shortest (green) to longest (red). (**C, D**) As the recurrence times are correlated, we find that individuals with a shorter time to the first recurrence (green) also have a shorter time from first to second recurrence (the data are shown in bolder lines and darker colors). (**C**) The temporal heterogeneity model cannot capture this feature in the data. The simulations show that all individuals have a similar time from first to second recurrence, regardless of the time to the first recurrence (simulated data are shown in thinner lines and lighter colors). (**D**) In the data simulated under the population heterogeneity model, however, the first recurrence time is predictive of the second recurrence time and this correlation agrees well with the data. The simulated data for chloroquine treatment compared with the Thailand-Myanmar data are shown in ***Supplementary Fig. S12***.

### Spearman correlation between time to first recurrence and time from first to second recurrence

### Both population and temporal heterogeneity contribute to relapse rates

The analysis to this point attempted to compare the temporal and population models to identify which mechanism better explained the observed patterns of relapse in these data. However, both factors may also operate concurrently. To investigate this, we investigated whether adding temporal heterogeneity to the model of population heterogeneity would improve the fit to the first and second recurrence data. Adding temporal heterogeneity in relapse risk significantly improved the fit compared to population heterogeneity model alone (AIC difference of 22, likelihood-ratio test with p-value < 0.0001, see ***Supplementary Fig. S11***). Together this indicates that in addition to population heterogeneity in risk of relapse, there is evidence for temporal changes in risk of relapse.

### Heterogeneity in exposure to *P vivax* infection may contribute to heterogeneity in relapse risk

One mechanism for population variation in hypnozoite number and relapse risk is if the frequency of a new infectious bite is variable across the population. If a single individual is more exposed to infection, then they may be more likely to have a larger hypnozoite reservoir [6, 26]. Since incidence rates for each individual were not easily discerned from these data, we instead considered whether higher transmission in a community led to higher rates of relapse. We did not have the necessary data to explore this question in the Thailand-Myanmar studies, but in the Papua New Guinea study, we could investigate variation in the recurrence risk by village by allowing both the risk of new mosquito-borne infections and the risk of relapses to vary between villages [4]. The 5 villages included in the Papua New Guinea trial had distinct transmission intensities [4]. Therefore, fitting our model of population heterogeneity to Papua New Guinea data stratified by village, we estimated the average risk of new infection and the average rate of relapse within each village. We found a weak (non-significant) association between the risk of new infections and the median relapse risk within each village (***Figure 5F***).

**Figure 5.**
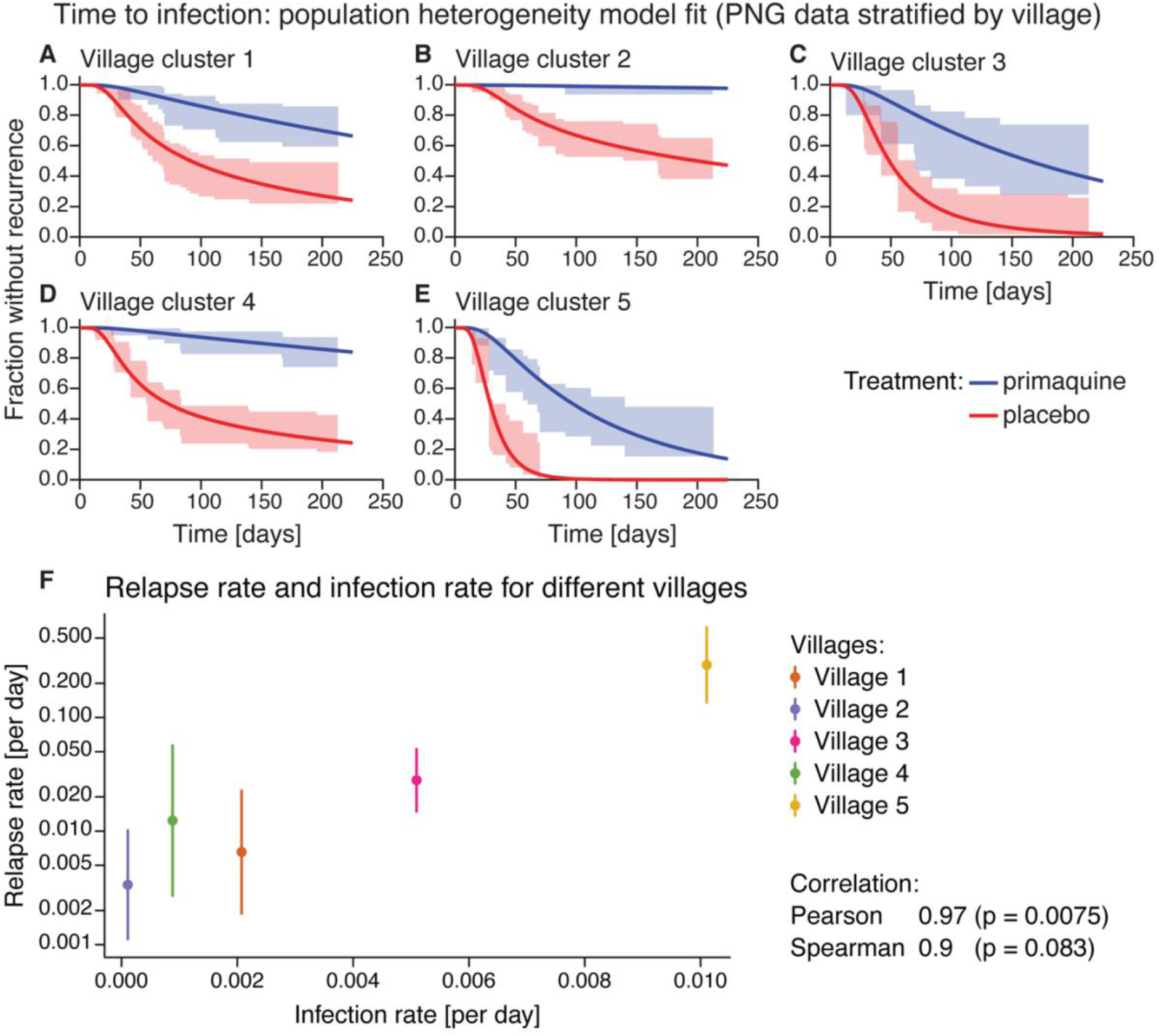
Time to infection population heterogeneity model fit and relapse rate vs infection rate for different villages in the Papua New Guinea data. (**A-E**) Model fit of the population heterogeneity model to the time to infection data from Papua New Guinea stratified by village. All villages were fit simultaneously with the same drug washout time distribution, the rate of new infections and relapses was allowed to vary between villages. The lines indicate the model fit and the shaded area the 95% confidence region from the data. (**F**) Relapse rate and infection rate for different villages. For each village, the median relapse rate (dot) and interquartile range (vertical line) of the relapse rate distribution from the population heterogeneity model fit is plotted against the infection rate. The Pearson and Spearman correlation between the log-transformed median relapse rate and the infection rate are 0.97 and 0.9, respectively, with p-values of 0.0075 and 0.083, respectively. For model fits to the Papua New Guinea data by village using the other models see ***Supplementary Fig. S3*** and ***Fig. S4***.

## Discussion

Here we provide strong evidence of variation in individuals propensity to experience relapses and demonstrate that population heterogeneity is a main driver of the overall pattern of recurrences observed in some endemic settings. Although our model of population heterogeneity did not explicitly incorporate mechanisms that can cause this population heterogeneity in relapse risk, some previous observations highlight mechanisms that are expected to give rise to such heterogeneity between individuals [14]. In monkey studies, the sporozoite inoculation size has been found to influence relapse frequency [20]. Indeed, natural sporozoite inoculum sizes are highly variable [27, 28]. In addition, differences in the hypnozoite reservoir between individuals could arise because of differences in individuals’ exposure to inoculation by infected mosquitoes [14, 22, 29]. Our analysis of the Papua New Guinea data stratified by village indicates that the average relapse risk in the community increases with increasing infection risk (***Figure 5***). Thus, spatial heterogeneity in the infection risk is likely an important determinant of relapse heterogeneity. Regional variation is also expected due to different parasite variants, with ‘temperate’ and ‘tropical’ strains of *P vivax* known to have different relapse rates and, in some regions, different strains appear to be present [6, 30, 31]. Other known factors that may contribute to population heterogeneity in *P vivax* malaria are immunity, age, heterogeneity in transmission within a community [14, 29, 32]. In particular, with higher blood-stage immunity, more asymptomatic, sub-microscopic infections may be expected which may be missed in clinical trials and cohort studies. This may present as some individuals having an apparent low relapse rate. Since spatial heterogeneity is likely an important determinant of relapse heterogeneity, we hypothesize that variable inoculum size and biting intensity between individuals explains much of the population heterogeneity in risk of relapse observed in these settings.

In addition to population heterogeneity, we found evidence that risk of relapse varies with time. This again is consistent with and follows from observations of temporal factors that have been observed to be associated with relapse risk, such as febrile illness [10] and even time since last infectious bite, since the reservoir of hypnozoites may deplete with each relapse [6].

Relapse heterogeneity can influence our understanding of the epidemiology of *P vivax* malaria. For example, the fraction of blood-stage infections that are relapses is often estimated to demonstrate the importance of relapse prevention and treatment with radical cure [7, 9]. However, estimates are often calculated assuming a constant relapse rate across the population and differ across studies [4, 7, 8]. In part this is due to whether the study has a clinical or epidemiological focus. For example, the studies considered here from the Thailand-Myanmar border region recruited individuals after having symptomatic *P vivax* malaria and thus inherently select for those with the highest risk of recurrences in the population [21, 25]. In contrast, the study from Papua New Guinea recruited individuals regardless of whether they had a *P vivax* episode at enrolment [4]. Indeed, Robinson et al., who enrolled and treated children regardless of their infection status, found that approximately 80% of blood-stage infections were relapses [4], compared with estimates of >90% from studies that enrolled *P vivax*-infected individuals [7, 8].

A factor that has not been included in our models of recurrence is the possibility of undetectable submicroscopic infections sustained by other hidden reservoirs of infection [33, 34]. Rather, we have only considered detectable infection and relapse. The prevalence of submicroscopic *P vivax* infection is often higher than the prevalence of microscopic infections [35] and may be an important factor for transmission of *P vivax*. Robinson et al. found that age was not significantly associated with the risk of the first *P vivax* blood-stage infection diagnosed with qPCR but the risk of microscopy-detectable *P vivax* infections decreased significantly with age [4]. Thus, our models may be biased towards younger and less immune individuals who are more likely to have microscopy-detectable infections [36, 37].

In this work, we found that population heterogeneity can capture observed patterns of the first and second recurrence times as well as the correlation between the time to first and second recurrence. This suggests that population heterogeneity plays an important role in the overall epidemiology of *P vivax* recurrence within a given year.

## Materials and Methods

### Data: Papua New Guinea data

The Papua New Guinea data set contains the data from a randomized placebo-controlled trial of liver-stage treatment using primaquine. The data was made publicly available by Robinson et al. [4] (where the details of the study can be found). The trial was conducted between August 2009 and 20 May 2010 in 5 village clusters in the Maprik District, East Sepik Province, Papua New Guinea. Children were enrolled and treated for blood-stage infections with chloroquine or artemether-lumefantrine and with either primaquine for liver-stage infections or with a placebo. After initial treatment, there were fortnightly active surveillance visits for 32 weeks and passive surveillance throughout the trial. The data contains (amongst others) the time from enrollment to *P vivax* infection by PCR and microscopy from 504 children as well as the children’s age (between 4 and 10 years), village cluster, and their treatment (placebo or primaquine). For the model fitting, we used the time to *P vivax* infection by PCR data and the village cluster information.

### Data: Thailand-Myanmar border region data

We consider the combined data from two randomized trials conducted in the Thailand-Myanmar border region, the VivaX History trial (VHX) [25] and the Best Primaquine Dose trial (BPD) [21] as published by Taylor et al. and made available online [8]. Details of the studies have been published previously [8, 21, 25]. In short, the VHX study was conducted between May 2010 and October 2012, it included 644 patients who were enrolled with uncomplicated *P vivax* malaria and randomized to either artesunate monotherapy, chloroquine monotherapy, or chloroquine with high dose primaquine. The BPD study was conducted between February 2012 and July 2014, it included 655 patients with symptomatic *P vivax* malaria, randomized to a primaquine (at one of two different doses) with either chloroquine or dihydroartemisinin-piperaquine (***Supplementary Table S9***). In both studies, infections were diagnosed using a malaria smear and antimalarial treatment was allocated based on a factorial design. In the VHX study, patients were excluded from primaquine treatment if they were found to be glucose-6-phosphate dehydrogenase (G6PD) deficient and in the BPD study G6PD deficiency was an exclusion criterion. Individuals were followed up for one year from enrolment and recurring *P vivax* infections were treated with the same antimalarial treatment as the first *P vivax* infection (VHX study) or with the standard chloroquine and primaquine regimen (BPD study). The data includes a patient id, episode number, antimalarial treatment, time from last event to current event, censored variable, study, and overall follow-up time.

One individual was excluded from the data as the data indicates censoring at the time of enrolment. This leaves 1298 individuals (446 patients who received blood-stage treatment and 852 patients who received both primaquine and blood-stage treatment). Data was analyzed in R (version 3.6.0) [38] using the survival [39, 40] and survminer [41] packages.

## Models

We considered four different mathematical models for the risk of relapse and recurrence in the cohorts. All models include a prophylactic effect of the antimalarial before drug washout and a constant rate of new infections. Patients are protected at enrolment due to the prophylactic effect of the antimalarial treatment, the time to drug washout is assumed to be lognormal distributed, and after drug washout, individuals become susceptible to new infection and relapses (see ***Supplementary Fig. S14*** for a model scheme). The models differ in the construction of the relapse rate.

### Model 1: constant relapse rate

In model 1, the recurrence rate is the sum of the constant rate of new infections and the constant relapse rate. Thus, the dynamics of the fraction of susceptible individuals *S*(*t*)at time *t* is given by:

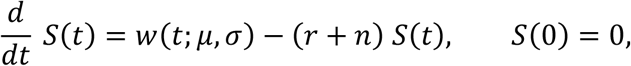

where *w*(*t*:*μ,σ*) is the probability density function of the lognormal distribution with parameters *μ* and *σ, r* is the constant relapse rate, and *n* is the constant rate of new infections.

### Model 2: temporal heterogeneity

Model 2 considers a relapse rate that is time dependent. Since our data analysis suggests that the relapse rate decreases after an initial peak (***Figure 1***), we chose a decreasing relapse rate such that the prophylactic effect of the antimalarial treatment in combination with the decreasing relapse rate can capture the observed change in the relapse rate (***Figyure 1***). We assume that there is an initial relapse rate (*I*) and that the relapse rate decreases exponentially over time (with rate *d*). Thus, the relapse rate is given by *r*(*t*) = *Ie*^−*dt*^ and the dynamics for the fraction of susceptible individuals at time *S(t)*is given by:

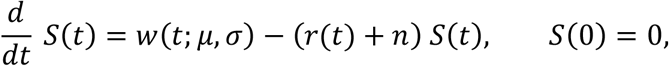

where *w*(*t*:*μ,σ*)is the probability density function of the lognormal distribution with parameters and *μ* and *σ, r*(*t*) = *Ie*^−*dt*^ is the time-dependent relapse rate, and *n* is the constant rate of new infections.

### Model 3: population heterogeneity

In model 3, the relapse rate is constant but drawn from a lognormal distribution to model population heterogeneity. To simplify the numerical computations, we group the population in *k* different risk groups of equal size. The relapse rate for each risk group is the median relapse rate for this group that is computed using the percentiles of the lognormal distribution of relapse rates (see ***Supplementary Fig. S15***). Thus, model 3 is given by:

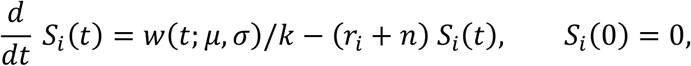

where *S* _*i*_ (*t*) is the fraction of susceptible individuals that are in risk group *i*(*i*∈{1,2. … .*k*}) at time *t,k* is the number of relapse risk groups, *r*_*i*_*(t) = I*_*i*_*e*^-dt^ is the median relapse rate of group *ri*, and *n* is the constant mosquito-borne infection rate. The overall fraction of susceptible individuals at time *t,S* (*t*), is the sum of the fractions of all susceptible individuals in the different risk groups:

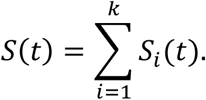

### Model 4: temporal and population heterogeneity

Finally, we considered a model that considers both temporal and population heterogeneity and is a combination and extension of models 2 and 3. As for model 3, we group the population in *k* different relapse risk groups of equal size and as for model 2 this relapse risk decreases over time, i.e., *r*_*i*_(*t*) = *I_i_e*^−*dt*^ where *I_i_* is the initial relapse risk for relapse risk group. The initial relapse risk (*I*_*i*_) is the median relapse rate for this group that is computed using the percentiles of the lognormal distribution of relapse rates (see ***Supplementary Fig. S15)***. Model 4 is given by:

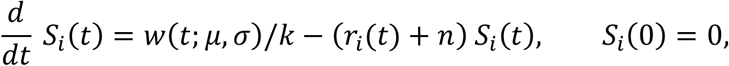

where *S* _*i*_ *(t)* is the fraction of susceptible individuals that are in risk group *i*(*i* ∈{1,2,…*k*}) at time *t,k* is the number of relapse risk groups, *r*_*i*_*(t) = I*_*i*_*e*^-dt^ is the time-dependent relapse rate of group *i*, and *n* is the constant mosquito-borne infection rate. As for model 3, the overall fraction of susceptible individuals at time *t,S(t)*, is the sum of the fractions of susceptible individuals in the different risk groups.

To fit the models not only to the first recurrence after enrolment but to the first and second recurrence, we extended the models to take two recurrences into account by adding additional compartments to the model (see Supplementary methods, ***Supplementary Fig. S16***). After a susceptible individual has a *P vivax* recurrence, the patient is again protected with the same drug washout time distribution as at enrolment because patients are treated with the same antimalarials at each recurrence. As for the first recurrence, patients become susceptible to mosquito-borne infections and relapses after drug washout.

For all models and all simulations shown, we assume that the drug washout time depends on the antimalarial treatment, the rate of new infections differs between the two different studies in the Thailand-Myanmar data as incidence rates changed between the time of the two studies [8] (***Supplementary Table S23*** and ***Fig. S17***), and we assumed that patients treated with primaquine have no relapses, i.e., we assume they are subject only to new infections (as previously; see Model fitting below and Supplement).

### Model fitting to recurrence times

Each model was implemented as an ODE or a system of ODEs and solved numerically using the function ode15s in MATLAB [42]. We constructed a likelihood function using this numerical solution to fit our models to the recurrence time data (including censored time intervals) as described below and obtain maximum likelihood estimates for the model parameters.

From the numerical solution of the model equations, we obtained the fraction of uninfected individuals at time *t, U*(*t*), as the sum of the fraction of individuals who are still protected and the fraction of susceptible individuals:

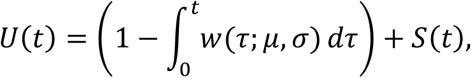

where *w(t; μ σ)*is the probability density function of the lognormal distribution of drug washout times with parameters *μ* and *σ*. We interpret *U(t)* as the probability of remaining uninfected until time *t*. The probability of having an infection at the follow-up visit on day *t* (*G*(*t*)) is then the probability of being uninfected on the previous follow-up visit but infected on day *t*, i.e.,

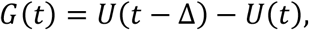

where *t*– Δ is the time of the last follow-up visit before day *t*.

Using *U(t)*, the probability of being uninfected until time *t*, and *G(t)*, the probability of having an infection on day *t*, we used the following loglikelihood function to fit the models to the first recurrence times:

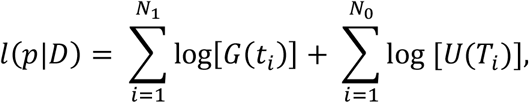

where *p* are the parameters of the model (see Supplementary methods for the parameters of the different models), *D* is the data, i.e., the recurrence times, *N*_0_ and *N*_1_ are the numbers of individuals with zero and at least one recurrence, respectively, *U*(*t*) is the probability of being uninfected until time *t, t*_*i*_ is the time of the first recurrence of individual *i, G*(*t*)is the probability of having an infection at the follow-up visit on day *t*, and *T*_*i*_ is the overall follow-up time of individual *i* (for censored time intervals).

For fitting the data to the first and second recurrence time simultaneously (in the Thailand-Myanmar data), we constructed a different loglikelihood function. We use both the first and the second recurrence information (as well as censoring times) from the data in our likelihood function and also incorporate the different relapse risk groups in models 3 and 4 to allow for heterogeneity in relapse rates across the population. The loglikelihood function for the parameter set *p* and the data *D* is given by:

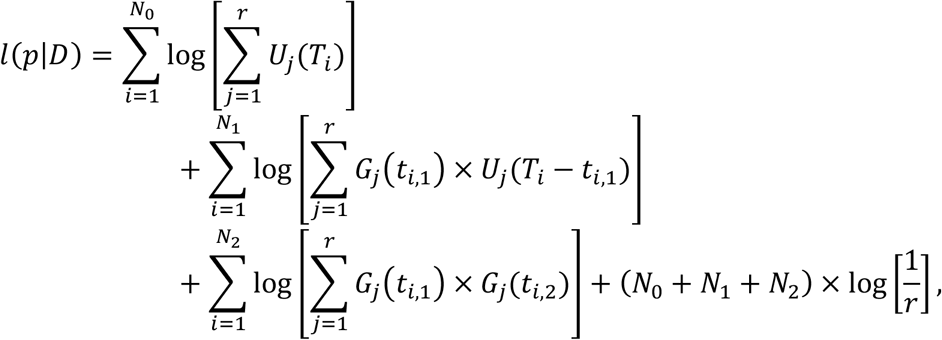

where *N*_0_, *N*_1_, and *N*_2_ are the numbers of individuals with 0, 1, or at least 2 recurrences, respectively, is the number of risk groups, *U*_*j*_(t) is the probability of remaining uninfected to time *t* for individuals in risk group *j,T*_*i*_ is the follow-up time of individual *i* (i.e., the time of censoring), *G*_*j*_ *(t)* is the probability of having an infection on the follow-up visit on day *t* for individuals in risk group *j*, and *t* _*i*,1_, *t* _*i*, 2_, are the times from the start of the study to the first recurrence and from the first to the second recurrence of individual *i*, respectively. Note this loglikelihood function can be simplified for models 1 and 2 as there is only one risk group in these models (see Supplementary methods for the derivation and more details on the loglikelihood function).

The model was fit by minimizing the negative loglikelihood function with the function fmincon in MATLAB [42]. We did 100 minimizations of the negative loglikelihood function with random initial parameter values to find the Maximum Likelihood Estimates (MLEs) for the parameters. Additionally, when fitting the extended model to the first and second recurrence data, we used the parameters of the best fit to the first recurrence times as initial parameter values. Confidence intervals were computed using bootstrapping and the percentile method (see Supplementary methods).

We fit models 1 to 3 using one mosquito-borne infection rate for the Papua New Guinea data and two different infection rates for the two studies in the Thailand-Myanmar data (***Supplementary Table S23*** and ***Fig. S17***). We also assessed the importance of assumptions regarding the follow-up schemes (***Supplementary Table S24, Fig. S18, Fig. S19***, and ***Fig. S20***), different numbers of relapse risk groups in model 3 (***Supplementary Table S25*** and ***Fig. S21***), and the relapse risk distribution in the population heterogeneity model (***Supplementary Table S26, Fig. S22***, and ***Fig. S23***) in the Thailand-Myanmar data. For the final model comparisons and simulations both for fitting to the first recurrence and for fitting to the first and second recurrence simultaneously, we chose two different infection rates, daily follow-ups, and 10 relapse risk groups for all models.

### Simulations of the models

To compare the model predictions of the associations in time to first relapse and time to second relapse with the Thailand-Myanmar data, we used models 2 and 3 to simulate data. We used the MLEs of the parameters (fit to the first and second recurrence times in the Thailand-Myanmar data) to simulate a population of individuals treated with artesunate (n=1000) or chloroquine (n=1000) monotherapy. Each individual was simulated by randomly drawing a drug washout time and recurrence times from the corresponding distribution parameterized during model fitting (***Supplementary Table S19*** and ***Table S20***). The drug washout time follows a lognormal distribution and the infection and relapse rates follow an exponential distribution. In the population heterogeneity model (model 3), each individual has a relapse rate that is drawn from a lognormal distribution and each individual’s relapse rate remains constant throughout the simulation. These simulations of 1000 individuals for models 2 and 3 and artesunate or chloroquine treatment were repeated 1000 times. Simulated individuals were censored after 365 days. We analyzed the simulated data in the same way as the original data and compared the results visually (***Figure 4*** and ***Supplementary Fig. S12)***. For more details on the model simulations see the Supplementary methods.

## Data Availability

The multiple recurrence data has been previous published and made available by Taylor et al. [8] in an online repository. The data from Papua New Guinea was published by Robinson et al. [4] and is available in an online repository.

https://github.com/jwatowatson/RecurrentVivax/blob/master/RData/TimingModel/Combined_Time_Event.RData

https://datadryad.org/stash/dataset/doi:10.5061/dryad.m1n03

## Acknowledgements

This work would not have been possible without the published data from [4] and from two studies by Chu et al. [21, 25] (made publicly available by [8]). We wish to thank the original study teams and participants for collecting this data and making it available for further comparative study and modelling.

This work was funded by the Australian Research Council (ARC) (grants DP120100064 & DP180103875 (to DSK, MPD, DC) and DP200100747 (to JAF)) and the National Health and Medical Research Council (NHMRC) of Australia [grants 1082022 (to MPD, DC), 1173528 (to DC), 1141921 (to DSK), 1080001 and 1173027 (to MPD) and 1135820 (to NMA)]. LJR is supported by NHMRC Career Development Fellowship (GNT1161627). The authors have no conflicts of interest to declare.

The authors thank Nicholas White and François Nosten for their integral roles in the two studies by [21, 25] and the work by [8] that provided data essential for this study and their feedback that helped improve the manuscript. The authors also thank Steffen Docken for helpful discussions.

## Author contributions

ES, DC, AIA, MPD, and DSK developed the mathematical models. ES fitted the mathematical models and analyzed the data. TES provided statistics expertise. ES, MPD, and DSK wrote the manuscript. SM, JAF, NMA, CSC, JW, and TES helped prepare the manuscript. All authors read and approved the final manuscript.

## Data and Code availability

The code will be made publicly available on GitHub upon publication.

