## Supplementary Information for "Risk of *Plasmodium vivax* recurrences follows a 30-70 rule and indicates relapse heterogeneity in the population"

Eva Stadler[1], Deborah Cromer[1], Somya Mehra[2], Adeshina I Adekunle[1,3], Jennifer A Flegg[2], Nicholas M Anstey[4], James A Watson[5,6], Cindy S Chu[5,7], Ivo Mueller[8,9], Leanne J Robinson[8,9,10,11], Timothy E Schlub[1,12], Miles P Davenport[1], David S Khoury[1]*

[1] The Kirby Institute, UNSW Sydney, Sydney, Australia
[2] School of Mathematics and Statistics, University of Melbourne, Melbourne, Australia
[3] Australian Institute of Tropical Health and Medicine, James Cook University, Townsville, Australia
[4] Menzies School of Health Research, Darwin University, Darwin, Australia
[5] Centre for Tropical Medicine and Global Health, Nuffield Department of Medicine, University of Oxford, United Kingdom
[6] Mahidol-Oxford Tropical Medicine Research Unit, Faculty of Tropical Medicine, Mahidol University, Bangkok, Thailand
[7] Shoklo Malaria Research Unit, Mahidol-Oxford Tropical Medicine Research Unit, Faculty of Tropical Medicine, Mahidol University, Mae Sot, Thailand
[8] Population Health & Immunity Division, Walter and Eliza Hall Institute of Medical Research, Parkville, Australia
[9] Department of Medical Biology, University of Melbourne, Melbourne, Australia
[10] Burnet Institute, Melbourne, Victoria, Australia
[11] PNG Institute of Medical Research, Madang, Papua New Guinea
[12] Sydney School of Public Health, Faculty of Medicine and Health, The University of Sydney, Sydney, Australia

### Additional tables

**Parameters of model 1: constant relapse rate (PNG data)**

| Parameter | MLE | 95% CI |
|---|---|---|
| Mean of the logarithmic values of the drug washout time distribution | 3.97 | [3.72,4.23] |
| Standard deviation of the logarithmic values of the drug washout time distribution | 1.81 | [1.47,2.24] |
| Recurrence rate for patients treated only for blood-stage infections [per day] | 0.044 | [0.035,0.056] |
| Recurrence rate for patients treated with primaquine [per day] | 0.0035 | [0.0025,0.0050] |

**Table S1** Maximum likelihood estimates of the parameters of the first model fit to the first recurrence time in the PNG data. Abbreviations: MLE Maximum Likelihood Estimate, CI Confidence Interval.

**Parameters of model 2: temporal heterogeneity (PNG data)**

| Parameter | MLE | 95% CI |
|---|---|---|
| Mean of the logarithmic values of the drug washout time distribution | 3.01 | [2.75, 3.24] |
| Standard deviation of the logarithmic values of the drug washout time distribution | 0.46 | [0.36,0.54] |
| Rate of new infections [per day] | 0.0019 | [0.0015,0.0024] |
| Initial relapse rate of patients [per day] | 0.033 | [0.020,0.063] |
| Exponential decay rate of the relapse rate [per day] | 0.022 | [0.015,0.030] |

**Table S2** Maximum likelihood estimates of the parameters of the second model fit to the first recurrence time in the PNG data. Abbreviations: MLE Maximum Likelihood Estimate, CI Confidence Interval.

**Parameters of model 3: population heterogeneity (PNG data)**

| Parameter | MLE | 95% CI |
|---|---|---|
| Mean of the logarithmic values of the drug washout time distribution | 3.11 | [2.83,3.65] |
| Standard deviation of the logarithmic values of the drug washout time distribution | 0.48 | [0.38,0.55] |
| Rate of new infections [per day] | 0.0019 | [0.0015,0.0025] |
| Mean of the logarithmic values of the relapse rate distribution | -4.73 | [-5.29,45.39] |
| Standard deviation of the logarithmic values of the relapse rate distribution | 2.26 | [1.48,389.9] |

**Table S3** Maximum likelihood estimates of the parameters of the third model fit to the first recurrence time in the PNG data. Abbreviations: MLE Maximum Likelihood Estimate, CI Confidence Interval.

**Parameters of model 4: temporal and population heterogeneity (PNG data)**

| Parameter | MLE | 95% CI |
|---|---|---|
| Mean of the logarithmic values of the drug washout time distribution | 3.01 | [2.78,3.23] |
| Standard deviation of the logarithmic values of the drug washout time distribution | 0.46 | [0.36,0.54] |
| Rate of new infections [per day] | 0.0019 | [0.0015,0.0024] |
| Mean of the logarithmic values of the initial relapse rate distribution | -3.40 | [-3.87,-2.84] |
| Standard deviation of the logarithmic values of the initial relapse rate distribution | 0.0040 | [0.0018,0.11] |
| Exponential decay rate of the relapse rate [per day] | 0.022 | [0.015,0.030] |

**Table S4** Maximum likelihood estimates of the parameters of the third model fit to the first recurrence time in the PNG data. Abbreviations: MLE Maximum Likelihood Estimate, CI Confidence Interval.

**Parameters of model 1: constant relapse rate (PNG data by village)**

| | Parameter | MLE | 95% CI |
|---|---|---|---|
| All villages | Mean of the logarithmic values of the drug washout time distribution | 3.7 | [2.65,4.00] |
| | Standard deviation of the logarithmic values of the drug washout time distribution | 1.44 | [0.28,1.71] |
| Village 1 | Recurrence rate for placebo group [per day] | 0.028 | [0.0055,0.047] |
| | Recurrence rate for primaquine group [per day] | 0.0031 | [0.0013,0.0050] |
| Village 2 | Recurrence rate for placebo group [per day] | 0.007 | [0.0031,0.014] |
| | Recurrence rate for primaquine group [per day] | 0.00014 | [0.00,0.0005] |
| Village 3 | Recurrence rate for placebo group [per day] | 0.055 | [0.018,0.0078] |
| | Recurrence rate for primaquine group [per day] | 0.0092 | [0.0026,0.19] |
| Village 4 | Recurrence rate for placebo group [per day] | 0.044 | [0.0068,0.066] |
| | Recurrence rate for primaquine group [per day] | 0.0013 | [0.005,0.0023] |
| Village 5 | Recurrence rate for placebo group [per day] | 0.12 | [0.054,0.19] |
| | Recurrence rate for primaquine group [per day] | 0.031 | [0.0062,0.066] |

**Table S5** Maximum likelihood estimates of the parameters of the constant relapse rate model fit to the first recurrence time in the PNG data with all villages fit simultaneously with the same drug washout time distribution. Abbreviations: MLE Maximum Likelihood Estimate, CI Confidence Interval.

**Parameters of model 2: temporal heterogeneity (PNG data by village)**

| | Parameter | MLE | 95% CI |
|---|---|---|---|
| All villages | Mean of the logarithmic values of the drug washout time distribution | 3.15 | [2.97,3.36] |
| | Standard deviation of the logarithmic values of the drug washout time distribution | 0.49 | [0.38,0.51] |
| Village 1 | Rate of new infections [per day] | 0.0020 | [0.0012,0.0031] |
| | Initial relapse rate of patients [per day] | 0.027 | [0.014,0.059] |
| | Exponential decay rate of the relapse rate [per day] | 0.019 | [0.011,0.030] |
| Village 2 | Rate of new infections [per day] | 0.00011 | [$1.42\times10^{-10}$,0.00036] |
| | Initial relapse rate of patients [per day] | 0.0090 | [0.0042,0.019] |
| | Exponential decay rate of the relapse rate [per day] | 0.0080 | [0.0011,0.017] |
| Village 3 | Rate of new infections [per day] | 0.0051 | [0.0024,0.0094] |
| | Initial relapse rate of patients [per day] | 0.043 | [0.020,0.11] |
| | Exponential decay rate of the relapse rate [per day] | 0.011 | [$3.06\times10^{-7}$,0.027] |
| Village 4 | Rate of new infections [per day] | 0.00091 | [0.00041,0.0015] |
| | Initial relapse rate of patients [per day] | 0.044 | [0.022,0.12] |
| | Exponential decay rate of the relapse rate [per day] | 0.022 | [0.013,0.037] |
| Village 5 | Rate of new infections [per day] | 0.010 | [0.0064,0.016] |
| | Initial relapse rate of patients [per day] | 0.24 | [0.068,12.59] |
| | Exponential decay rate of the relapse rate [per day] | 0.015 | [$4.20\times10^{-7}$,0.045] |

**Table S6** Maximum likelihood estimates of the parameters of the temporal heterogeneity model fit to the first recurrence time in the PNG data with all villages fit simultaneously with the same drug washout time distribution. Abbreviations: MLE Maximum Likelihood Estimate, CI Confidence Interval.

**Parameters of model 3: population heterogeneity (PNG data by village)**

| | Parameter | MLE | 95% CI |
|---|---|---|---|
| All villages | Mean of the logarithmic values of the drug washout time distribution | 3.2 | [2.91,3.45] |
| | Standard deviation of the logarithmic values of the drug washout time distribution | 0.49 | [0.38,0.54] |
| Village 1 | Rate of new infections [per day] | 0.0021 | [0.0012,0.0034] |
| | Mean of the logarithmic values of the relapse rate distribution | -5.03 | [-6.45,-4.18] |
| | Standard deviation of the logarithmic values of the relapse rate distribution | 1.85 | [1.04,4.34] |
| Village 2 | Rate of new infections [per day] | 0.00011 | [$3.37\times10^{-11}$,0.00036] |
| | Mean of the logarithmic values of the relapse rate distribution | -5.69 | [-6.68,-5.04] |
| | Standard deviation of the logarithmic values of the relapse rate distribution | 1.63 | [0.51,3.12] |
| Village 3 | Rate of new infections [per day] | 0.0051 | [0.0024,0.010] |
| | Mean of the logarithmic values of the relapse rate distribution | -3.57 | [-4.56,-2.53] |
| | Standard deviation of the logarithmic values of the relapse rate distribution | 0.94 | [0.0015,2.31] |
| Village 4 | Rate of new infections [per day] | 0.00088 | [0.00035,0.0015] |
| | Mean of the logarithmic values of the relapse rate distribution | -4.39 | [-5.41,-3.24] |
| | Standard deviation of the logarithmic values of the relapse rate distribution | 2.26 | [1.19,5.21] |
| Village 5 | Rate of new infections [per day] | 0.01 | [0.0060,0.016] |
| | Mean of the logarithmic values of the relapse rate distribution | -1.24 | [-2.83,$3.52\times10^{6}$] |
| | Standard deviation of the logarithmic values of the relapse rate distribution | 1.13 | [0.0011,$9.66\times10^{5}$] |

**Table S7** Maximum likelihood estimates of the parameters of the population heterogeneity model fit to the first recurrence time in the PNG data with all villages fit simultaneously with the same drug washout time distribution. Abbreviations: MLE Maximum Likelihood Estimate, CI Confidence Interval.

**Parameters of model 4: temporal and population heterogeneity (PNG data by village)**

| | Parameter | MLE | 95% CI |
|---|---|---|---|
| All villages | Mean of the logarithmic values of the drug washout time distribution | 3.18 | [2.94,3.40] |
| | Standard deviation of the logarithmic values of the drug washout time distribution | 0.51 | [0.39,0.56] |
| Village 1 | Rate of new infections [per day] | 0.0020 | [0.0011,0.0032] |
| | Mean of the logarithmic values of the initial relapse rate distribution | -3.56 | [-4.30,-2.82] |
| | Standard deviation of the logarithmic values of the initial relapse rate distribution | 0.015 | [0.0010,0.33] |
| | Exponential decay rate of the relapse rate [per day] | 0.019 | [0.010,0.030] |
| Village 2 | Rate of new infections [per day] | 0.00011 | [$1.53\times10^{-10}$,0.0004] |
| | Mean of the logarithmic values of the initial relapse rate distribution | -5.61 | [-6.30,-3.97] |
| | Standard deviation of the logarithmic values of the initial relapse rate distribution | 1.56 | [0.0073,2.26] |
| | Exponential decay rate of the relapse rate [per day] | 0.00074 | [$8.74\times10^{-8}$,0.017] |
| Village 3 | Rate of new infections [per day] | 0.0051 | [0.0022,0.010] |
| | Mean of the logarithmic values of the initial relapse rate distribution | -3.08 | [-4.05,-2.06] |
| | Standard deviation of the logarithmic values of the initial relapse rate distribution | 0.038 | [0.0009,1.11] |
| | Exponential decay rate of the relapse rate [per day] | 0.011 | [$1.49\times10^{-7}$,0.028] |
| Village 4 | Rate of new infections [per day] | 0.00092 | [0.0004,0.0016] |
| | Mean of the logarithmic values of the initial relapse rate distribution | -3.07 | [-3.97,-2.24] |
| | Standard deviation of the logarithmic values of the initial relapse rate distribution | 0.041 | [0.0010,1.59] |
| | Exponential decay rate of the relapse rate [per day] | 0.023 | [0.0080,0.037] |
| Village 5 | Rate of new infections [per day] | 0.010 | [0.0063,0.017] |
| | Mean of the logarithmic values of the initial relapse rate distribution | -1.36 | [-2.78,2.57] |
| | Standard deviation of the logarithmic values of the initial relapse rate distribution | 0.085 | [0.0024,2.02] |
| | Exponential decay rate of the relapse rate [per day] | 0.011 | [$1.68\times10^{-7}$,0.037] |

**Table S8** Maximum likelihood estimates of the parameters of the temporal and population heterogeneity model fit to the first recurrence time in the PNG data with all villages fit simultaneously with the same drug washout time distribution. Abbreviations: MLE Maximum Likelihood Estimate, CI Confidence Interval.

##### Antimalarial treatments by study in the Thailand-Myanmar data

| Study | **VivaX History study (VHX)** | **Best Primaquine Dose study (BPD)** |
|---|---|---|
| Treatment | Artesunate (AS)<br>Chloroquine (CHQ)<br>Chloroquine and primaquine (CHQ/PMQ) | Chloroquine and primaquine (CHQ/PMQ)<br>Dihydroartemisinin-piperaquine and primaquine (DP/PMQ) |

**Table S9** Antimalarial treatments by study in the Thailand-Myanmar data. In the VHX study, patients were treated with either artesunate (AS), chloroquine (CHQ), or chloroquine and primaquine (CHQ/PMQ). In the BPD study, patients were treated with chloroquine and primaquine (CHQ/PMQ) or with dihydroartemisinin-piperaquine and primaquine (DP/PMQ). These abbreviations are used through the supplement.

##### Contribution to recurrences by number of recurrences in the Thailand-Myanmar data

| **Number of recurrences** | **Number of individuals (%)** | | **Number of recurrences caused by individuals with _ recurrences (%)** | |
|---|---|---|---|---|
| 0 | 841 | (64.74%) | 0 | (0%) |
| 1 | 174 | (13.39%) | 174 | (12.07%) |
| 2 | 86 | (6.62%) | 172 | (11.94%) |
| 3 | 46 | (3.54%) | 138 | (9.58%) |
| 4 | 39 | (3.00%) | 156 | (10.83%) |
| 5 | 29 | (2.23%) | 145 | (10.06%) |
| 6 | 22 | (1.69%) | 132 | (9.16%) |
| 7 | 17 | (1.31%) | 119 | (8.26%) |
| 8 | 16 | (1.23%) | 128 | (8.88%) |
| 9 | 21 | (1.62%) | 189 | (13.12%) |
| 10 | 5 | (0.38%) | 50 | (3.47%) |
| 11 | 1 | (0.08%) | 11 | (0.76%) |
| 12 | 0 | (0%) | 0 | (0%) |
| 13 | 1 | (0.08%) | 13 | (0.90%) |
| 14 | 1 | (0.08%) | 14 | (0.97%) |

**Table S10** Contribution of individuals with different number of recurrences to the overall number of recurrences in the Thailand-Myanmar data. Data for both studies and all antimalarial treatments. This table shows, e.g., that the individuals with 3 or more recurrences are 15.2% of the population but they cause 75.99% of all recurrences. For a visualization of the contribution to recurrences see **Fig. S6**.

##### Spearman correlation between time to first recurrence and time from first to second recurrence in the Thailand-Myanmar data

| Data | All data (p-value) | | Excluding censored data (p-value) | |
|---|---|---|---|---|
| All data | 0.33 | (<0.0001*) | 0.63 | (<0.0001*) |
| VHX study | 0.39 | (<0.0001*) | 0.62 | (<0.0001*) |
| BPD study | -0.69 | (<0.0001*) | -0.54 | (0.09) |
| AS treated | 0.53 | (<0.0001*) | 0.59 | (<0.0001*) |
| CHQ treated | 0.31 | (<0.0001*) | 0.42 | (<0.0001*) |
| PMQ+ treated | -0.73 | (<0.0001*) | -0.63 | (0.01) |

**Table S11** Spearman correlation between time to first recurrence and time from first to second recurrence in the Thailand-Myanmar data. In the VHX study individuals were treated either with artesunate (AS), chloroquine (CHQ), or chloroquine and primaquine (PMQ+) and in the BPD study individuals were treated with chloroquine and primaquine or with dihydroartemisinin-piperaquine and primaquine (PMQ+). The asterisk (*) indicates that the exact p-value could not be computed in R due to ties.

**Spearman correlation between time to first recurrence and time from first to second recurrence with recurrence times restricted to 182 days**

| | Artesunate (p-value) | | Chloroquine (p-value) | |
|---|---|---|---|---|
| Model 1: constant relapse rate | 0.0008 | (0.52) | 0.0009 | (0.48) |
| Model 2: temporal heterogeneity | -0.001 | (0.39) | 0.002 | (0.079) |
| Model 3: population heterogeneity | 0.52 | (<0.0001) | 0.38 | (<0.0001) |
| Thailand-Myanmar data | 0.55 | (<0.0001) | 0.46 | (<0.0001) |

**Table S12** Spearman correlation between time to first recurrence and time from first to second recurrence in the simulated data and the TM data excluding censored data with recurrence times restricted to 182 days. All of the 1,000,000 simulated individuals who had at least two recurrences during the 1-year-simulation with both recurrences within the 182 days of the previous recurrence were used to compute the Spearman correlation. For the TM data, we show here the Spearman correlation for all individuals who had at least two known recurrences and each recurrence was within 182 days of the last known recurrence. The recurrence times were restricted to 182 days to avoid a bias in the second recurrence time due to the first recurrence time that is present if the sum of both recurrence times is restricted to be at most 365 days (e.g., if an individual has a long time to the first recurrence, then the time to the second recurrence is necessarily short, however if we restrict both recurrence times to 182 days, then the first recurrence time does not give any information about the second recurrence which may be at any time between 1 and 182 days after the first recurrence).

#### Cox regression on time from 1st to 2nd recurrence in the Thailand-Myanmar data

| | **AS treatment** | **CHQ treatment** |
|---|---|---|
| Hazard ratio | 0.987 | 0.989 |
| p-value | 0.0001 | 0.00004 |
| 95% CI | 0.9805-0.9937 | 0.9833-0.994 |

**Table S13** Cox regression of the time from first to second recurrence with time to first recurrence as a continuous variable for individuals treated with artesunate (AS) or chloroquine (CHQ) in the Thailand-Myanmar data. The hazard ratio shows that individuals with a longer time to the first recurrence have a lower risk of recurrence, i.e., they also have a longer time to their second recurrence.

**Parameters of model 1: constant relapse rate (for fitting to the first recurrence time in the Thailand-Myanmar data)**

| Parameter | MLE | 95% CI |
|---|---|---|
| Mean of the logarithmic values of the drug washout time distribution for AS | 3.26 | [3.08,3.46] |
| Standard deviation of the logarithmic values of the drug washout time distribution for AS | 1.69 | [1.33,2.15] |
| Mean of the logarithmic values of the drug washout time distribution for CHQ | 4.10 | [3.95,4.32] |
| Standard deviation of the logarithmic values of the drug washout time distribution for CHQ | 1.35 | [1.15,1.61] |
| Mean of the logarithmic values of the drug washout time distribution for DP | 3.88 | [3.86,6.27] |
| Standard deviation of the logarithmic values of the drug washout time distribution for DP | 0.057 | [0.052,4.34] |
| Recurrence rate for patients treated only for blood-stage infections (VHX study) [per day] | 0.0721 | [0.0606,0.0893] |
| Recurrence rate for patients treated with primaquine in the VHX study [per day] | 0.00106 | [0.0007,0.0015] |
| Recurrence rate for patients treated with primaquine in the BPD study [per day] | 0.00061 | [0.0005,0.001] |

**Table S14** Maximum likelihood estimates of the parameters of the first model fit to the first recurrence time in the Thailand-Myanmar data. Abbreviations: MLE Maximum Likelihood Estimate, CI Confidence Interval, AS artesunate treatment, CHQ chloroquine treatment, DP dihydroartemisinin-piperaquine treatment, VHX Vivax History study, BPD Best Primaquine Dose study.

**Parameters of model 2: temporal heterogeneity (for fitting to the first recurrence time in the Thailand-Myanmar data)**

| Parameter | MLE | 95% CI |
|---|---|---|
| Mean of the logarithmic values of the drug washout time distribution for AS | 2.81 | [2.74,2.88] |
| Standard deviation of the logarithmic values of the drug washout time distribution for AS | 0.26 | [0.15,0.34] |
| Mean of the logarithmic values of the drug washout time distribution for CHQ | 3.42 | [3.34,3.51] |
| Standard deviation of the logarithmic values of the drug washout time distribution for CHQ | 0.38 | [0.28,0.46] |
| Mean of the logarithmic values of the drug washout time distribution for DP | 3.89 | [3.86,3.94] |
| Standard deviation of the logarithmic values of the drug washout time distribution for DP | 0.057 | [0.053,0.063] |
| Rate of new infections in the VHX study [per day] | 0.00089 | [0.0007,0.0011] |
| Rate of new infections in the BPD study [per day] | 0.00053 | [0.0004,0.0006] |
| Initial relapse rate for patients treated with AS or CHQ only (VHX study) [per day] | 0.088 | [0.0682,0.12] |
| Exponential decay rate of the relapse rate [per day] | 0.029 | [0.025,0.035] |

**Table S15** Maximum likelihood estimates of the parameters of the second model fit to the first recurrence time in the Thailand-Myanmar data. Abbreviations: MLE Maximum Likelihood Estimate, CI Confidence Interval, AS artesunate treatment, CHQ chloroquine treatment, DP dihydroartemisinin-piperaquine treatment, VHX Vivax History study, BPD Best Primaquine Dose study.

**Parameters of model 3: population heterogeneity (for fitting to the first recurrence time in the Thailand-Myanmar data)**

| Parameter | MLE | 95% CI |
|---|---|---|
| Mean of the logarithmic values of the drug washout time distribution for AS | 3.11 | [3.05,3.18] |
| Standard deviation of the logarithmic values of the drug washout time distribution for AS | 0.28 | [0.21,0.34] |
| Mean of the logarithmic values of the drug washout time distribution for CHQ | 3.68 | [3.61,3.77] |
| Standard deviation of the logarithmic values of the drug washout time distribution for CHQ | 0.42 | [0.35,0.49] |
| Mean of the logarithmic values of the drug washout time distribution for DP | 3.90 | [1.88,4.76] |
| Standard deviation of the logarithmic values of the drug washout time distribution for DP | 0.063 | [0.057,20.42] |
| Rate of new infections in the VHX study [per day] | 0.00078 | [0.0006,0.001] |
| Rate of new infections in the BPD study [per day] | 0.00054 | [0.0005,0.0007] |
| Mean of the logarithmic values of the relapse rate distribution for AS and CHQ (VHX study) | -1.88 | [-2.66,-0.42] |
| Standard deviation of the logarithmic values of the relapse rate distribution for AS and CHQ (VHX study) | 5.05 | [3.79,7.50] |

**Table S16** Maximum likelihood estimates of the parameters of the third model fit to the first recurrence time in the Thailand-Myanmar data. Abbreviations: MLE Maximum Likelihood Estimate, CI Confidence Interval, AS artesunate treatment, CHQ chloroquine treatment, DP dihydroartemisinin-piperaquine treatment, VHX Vivax History study, BPD Best Primaquine Dose study.

**Parameters of model 4: temporal and population heterogeneity (for fitting to the first recurrence time in the Thailand-Myanmar data)**

| Parameter | MLE | 95% CI |
|---|---|---|
| Mean of the logarithmic values of the drug washout time distribution for AS | 3.09 | [3.03,3.14] |
| Standard deviation of the logarithmic values of the drug washout time distribution for AS | 0.27 | [0.21,0.36] |
| Mean of the logarithmic values of the drug washout time distribution for CHQ | 3.68 | [3.60,3.78] |
| Standard deviation of the logarithmic values of the drug washout time distribution for CHQ | 0.42 | [0.33,0.51] |
| Mean of the logarithmic values of the drug washout time distribution for DP | 3.87 | [3.84,3.95] |
| Standard deviation of the logarithmic values of the drug washout time distribution for DP | 0.062 | [0.056,0.068] |
| Rate of new infections in the VHX study [per day] | 0.00083 | [0.0006,0.0011] |
| Rate of new infections in the BPD study [per day] | 0.00055 | [0.0004,0.0007] |
| Mean of the logarithmic values of the initial relapse rate distribution for AS and CHQ (VHX study) | -1.60 | [-1.92,-1.41] |
| Standard deviation of the logarithmic values of the initial relapse rate distribution for AS and CHQ (VHX) | 3.93 | [3.67,4.10] |
| Exponential decay rate of the relapse rate [per day] | 0.0081 | [0.0051,0.013] |

**Table S17** Maximum likelihood estimates of the parameters of the fourth model fit to the first recurrence time in the Thailand-Myanmar data. Abbreviations: MLE Maximum Likelihood Estimate, CI Confidence Interval, AS artesunate treatment, CHQ chloroquine treatment, DP dihydroartemisinin-piperaquine treatment, VHX Vivax History study, BPD Best Primaquine Dose study.

**Parameters of model 1: constant relapse rate (Thailand-Myanmar data)**

| Parameter | MLE | 95% CI |
|---|---|---|
| Mean of the logarithmic values of the drug washout time distribution for AS | 3.44 | [3.29,3.62] |
| Standard deviation of the logarithmic values of the drug washout time distribution for AS | 1.61 | [1.40,1.87] |
| Mean of the logarithmic values of the drug washout time distribution for CHQ | 4.05 | [3.92,4.20] |
| Standard deviation of the logarithmic values of the drug washout time distribution for CHQ | 1.15 | [1.02,1.29] |
| Mean of the logarithmic values of the drug washout time distribution for DP | 3.89 | [2.07,5.81] |
| Standard deviation of the logarithmic values of the drug washout time distribution for DP | 0.057 | [0.052,16.64] |
| Recurrence rate for patients treated only for blood-stage infections (VHX study) [per day] | 0.081 | [0.072,0.095] |
| Recurrence rate for patients treated with primaquine in the VHX study [per day] | 0.0011 | [0.0007,0.0014] |
| Recurrence rate for patients treated with primaquine in the BPD study [per day] | 0.00065 | [0.0005,0.001] |

**Table S18** Estimates and 95% confidence intervals for the parameters of the first model fit simultaneously to the first and second recurrence time in the Thailand-Myanmar data. Abbreviations: MLE Maximum Likelihood Estimate, CI Confidence Interval, AS artesunate treatment, CHQ chloroquine treatment, DP dihydroartemisinin-piperaquine treatment, VHX Vivax History study, BPD Best Primaquine Dose study.

**Parameters of model 2: temporal heterogeneity (Thailand-Myanmar data)**

| Parameter | MLE | 95% CI |
|---|---|---|
| Mean of the logarithmic values of the drug washout time distribution for AS | 2.79 | [2.75,2.86] |
| Standard deviation of the logarithmic values of the drug washout time distribution for AS | 0.24 | [0.18,0.29] |
| Mean of the logarithmic values of the drug washout time distribution for CHQ | 3.38 | [3.33,3.45] |
| Standard deviation of the logarithmic values of the drug washout time distribution for CHQ | 0.31 | [0.26,0.36] |
| Mean of the logarithmic values of the drug washout time distribution for DP | 3.88 | [3.86,3.88] |
| Standard deviation of the logarithmic values of the drug washout time distribution for DP | 0.057 | [0.054,0.058] |
| Rate of new infections in the VHX study [per day] | 0.00097 | [0.0008,0.0012] |
| Rate of new infections in the BPD study [per day] | 0.00057 | [0.0005,0.0007] |
| Initial relapse rate for patients treated with AS or CHQ only (VHX study) [per day] | 0.070 | [0.061,0.089] |
| Exponential decay rate of the relapse rate [per day] | 0.025 | [0.022,0.029] |

**Table S19** Estimates and 95% confidence intervals for the parameters of the second model fit simultaneously to the first and second recurrence time in the Thailand-Myanmar data. Abbreviations: MLE Maximum Likelihood Estimate, CI Confidence Interval, AS artesunate treatment, CHQ chloroquine treatment, DP dihydroartemisinin-piperaquine treatment, PMQ primaquine treatment, VHX Vivax History study, BPD Best Primaquine Dose study.

**Parameters of model 3: population heterogeneity (Thailand-Myanmar data)**

| Parameter | MLE | 95% CI |
|---|---|---|
| Mean of the logarithmic values of the drug washout time distribution for AS | 2.95 | [2.90,3.00] |
| Standard deviation of the logarithmic values of the drug washout time distribution for AS | 0.24 | [0.18,0.29] |
| Mean of the logarithmic values of the drug washout time distribution for CHQ | 3.53 | [3.46,3.60] |
| Standard deviation of the logarithmic values of the drug washout time distribution for CHQ | 0.33 | [0.25,0.39] |
| Mean of the logarithmic values of the drug washout time distribution for DP | 3.88 | [-4.86,4.74] |
| Standard deviation of the logarithmic values of the drug washout time distribution for DP | 0.060 | [0.056,27.46] |
| Rate of new infections in the VHX study [per day] | 0.0008 | [0.0006,0.0011] |
| Rate of new infections in the BPD study [per day] | 0.0006 | [0.0005,0.0008] |
| Mean of the logarithmic values of the relapse rate distribution for AS and CHQ (VHX study) | -3.73 | [-4.03,-3.39] |
| Standard deviation of the logarithmic values of the relapse rate distribution for AS and CHQ (VHX study) | 2.80 | [2.39,3.26] |

**Table S20** Estimates and 95% confidence intervals for the parameters of the third model fit simultaneously to the first and second recurrence time in the Thailand-Myanmar data. Abbreviations: MLE Maximum Likelihood Estimate, CI Confidence Interval, AS artesunate treatment, CHQ chloroquine treatment, DP dihydroartemisinin-piperaquine treatment, PMQ primaquine treatment, VHX Vivax History study, BPD Best Primaquine Dose study.

**Parameters of model 4: temporal and population heterogeneity (Thailand-Myanmar data)**

| Parameter | MLE | 95% CI |
|---|---|---|
| Mean of the logarithmic values of the drug washout time distribution for AS | 2.93 | [2.88,2.98] |
| Standard deviation of the logarithmic values of the drug washout time distribution for AS | 0.24 | [0.17,0.24] |
| Mean of the logarithmic values of the drug washout time distribution for CHQ | 3.54 | [3.47,3.60] |
| Standard deviation of the logarithmic values of the drug washout time distribution for CHQ | 0.33 | [0.26,0.40] |
| Mean of the logarithmic values of the drug washout time distribution for DP | -10.23 | [-26.66,4.56] |
| Standard deviation of the logarithmic values of the drug washout time distribution for DP | 26.62 | [21.01,31.89] |
| Rate of new infections in the VHX study [per day] | 0.00089 | [0.0007,0.0011] |
| Rate of new infections in the BPD study [per day] | 0.00062 | [0.0005,0.0008] |
| Mean of the logarithmic values of the relapse rate distribution for AS and CHQ (VHX study) | -3.01 | [-3.33,-2.68] |
| Standard deviation of the logarithmic values of the relapse rate distribution for AS and CHQ (VHX study) | 1.86 | [1.53,2.24] |
| Exponential decay rate of the relapse rate [per day] | 0.012 | [0.0083,0.017] |

**Table S21** Estimates and 95% confidence intervals for the parameters of the fourth model fit simultaneously to the first and second recurrence time in the Thailand-Myanmar data. Abbreviations: MLE Maximum Likelihood Estimate, CI Confidence Interval, AS artesunate treatment, CHQ chloroquine treatment, DP dihydroartemisinin-piperaquine treatment, PMQ primaquine treatment, VHX Vivax History study, BPD Best Primaquine Dose study.

#### Additional figures

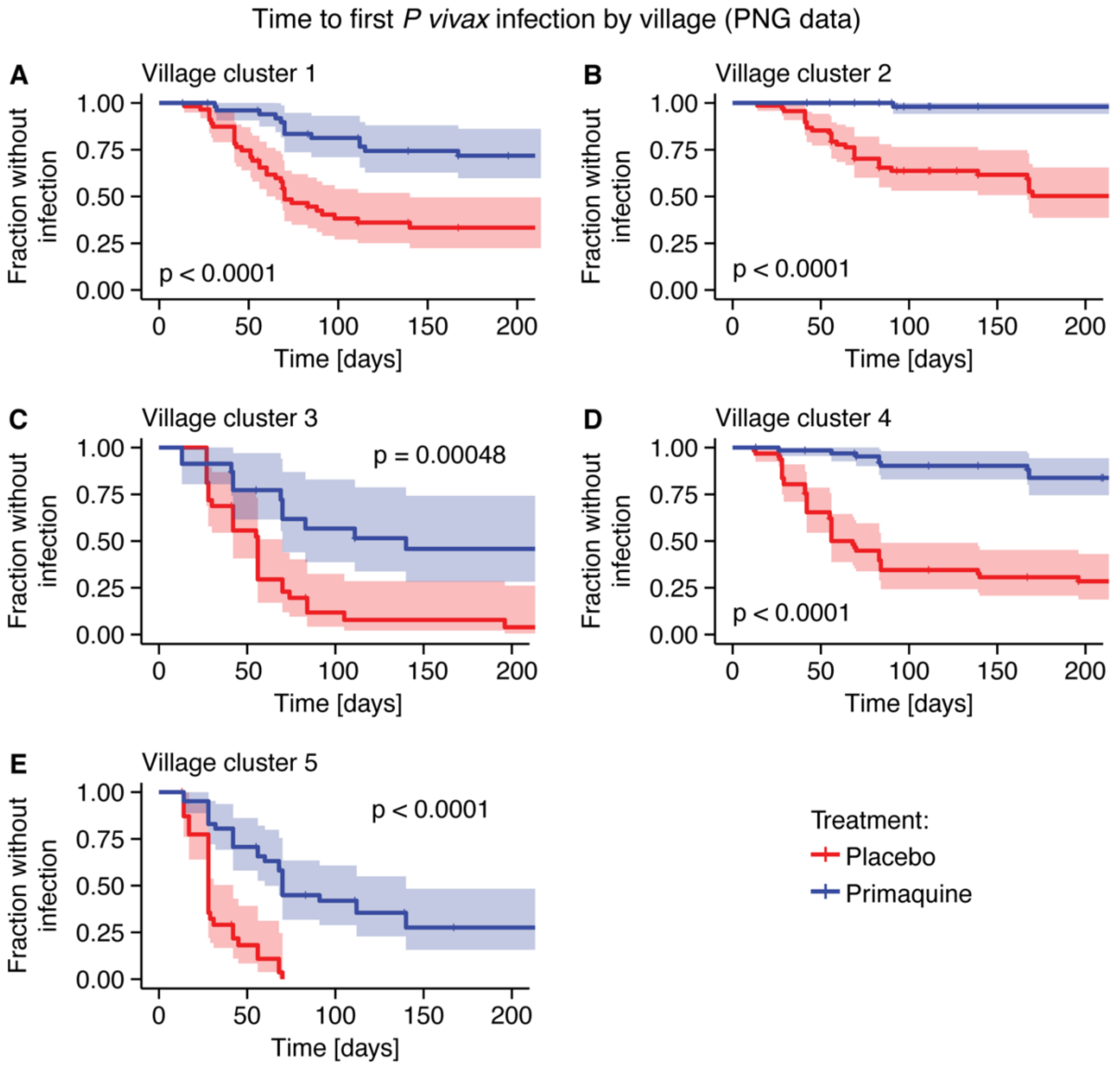


**Fig. S1** Time to first *P vivax* infection by village in the PNG data for patients treated with primaquine (blue) and a placebo (red). The placebo-treated patients have a higher infection risk in all villages (log-rank test with p-values < 0.001, see figures). Fits of the constant relapse risk, temporal heterogeneity, population heterogeneity, and temporal and population heterogeneity models all show different risks of new, mosquito-borne infections between villages with the lowest risk in village 2, followed by villages 4, 1, 3, and the highest risk in village 5 (see **Table S5**, **Table S6**, **Table S7**, and **Table S8**).

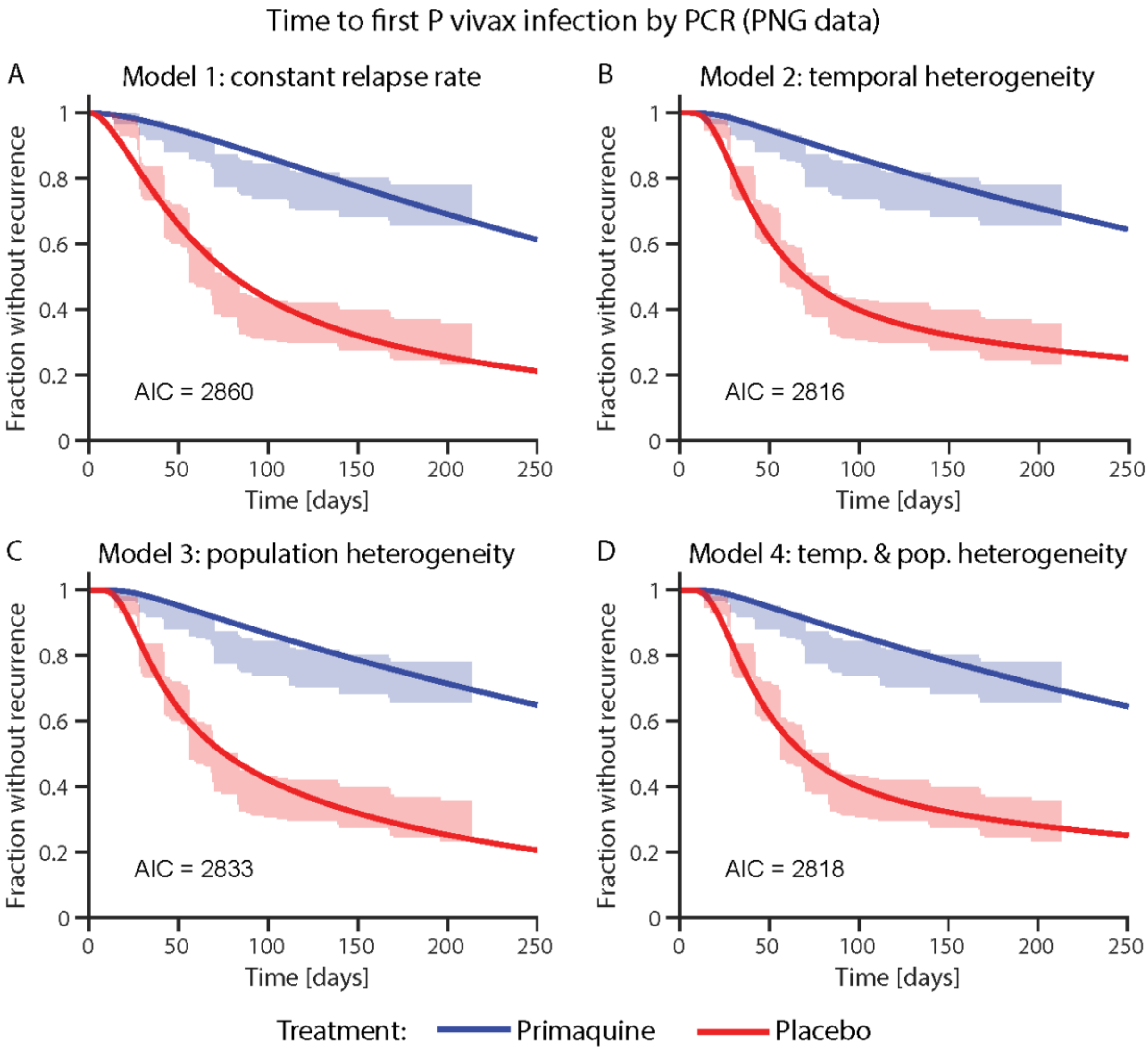


**Fig. S2** Model fit of the constant relapse rate, temporal heterogeneity, the population heterogeneity, and temporal and population heterogeneity model to the first recurrence time in the PNG data. The shaded areas are the 95% confidence regions from the data. For the parameters of the model fit see **Table S1**, **Table S2**, **Table S3**, and **Table S4**.

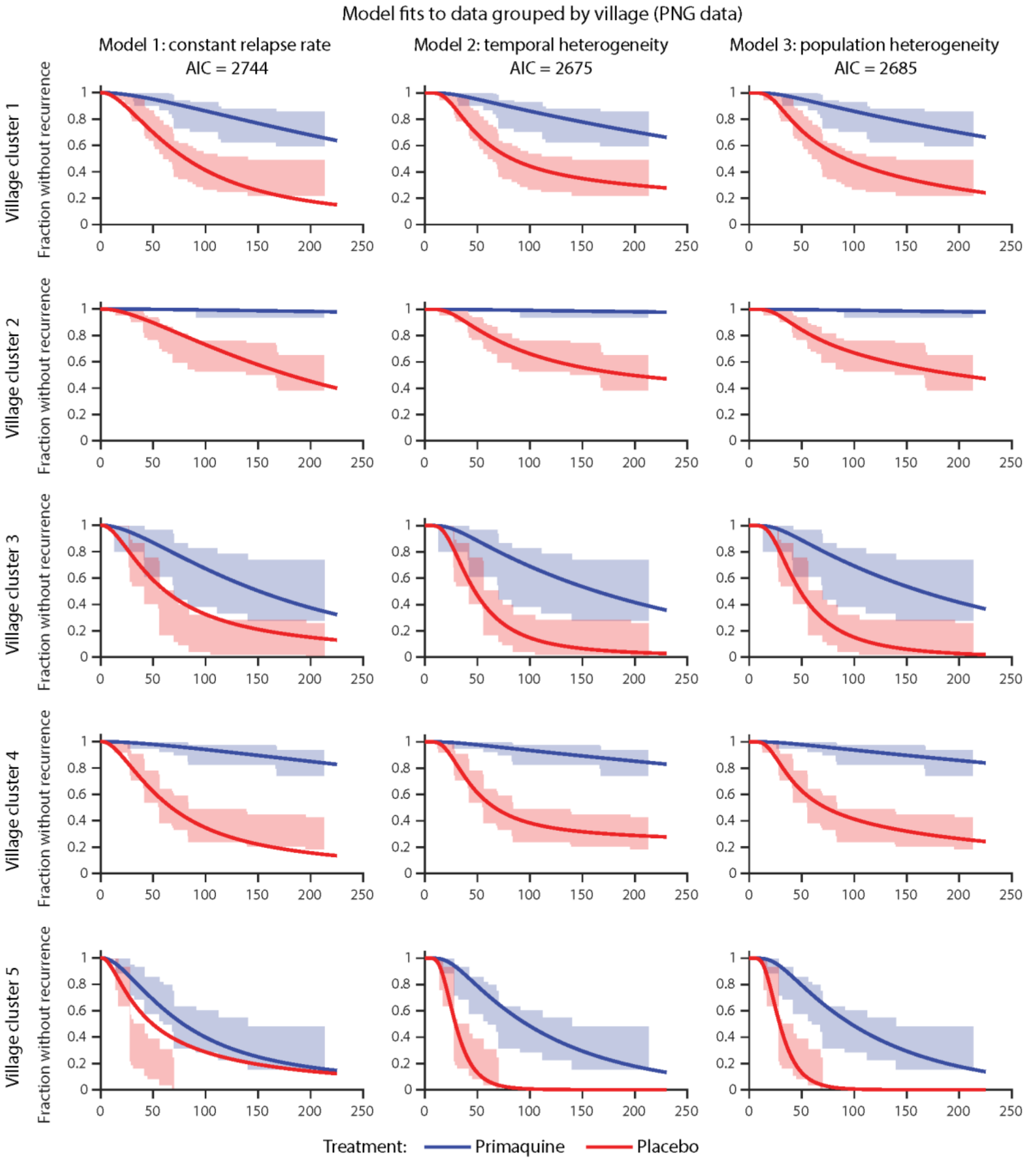


**Fig. S3** Fit of models 1 to 3 to the time to first *P vivax* infection by PCR in the PNG data grouped by village. All villages were fit simultaneously with the same drug washout time distribution, the rate of new infections and relapses were allowed to vary between villages. The lines indicate the model fit and the shaded area the 95% confidence region from the data. For the parameters of the model fit see **Table S5**, **Table S6**, and **Table S7**.

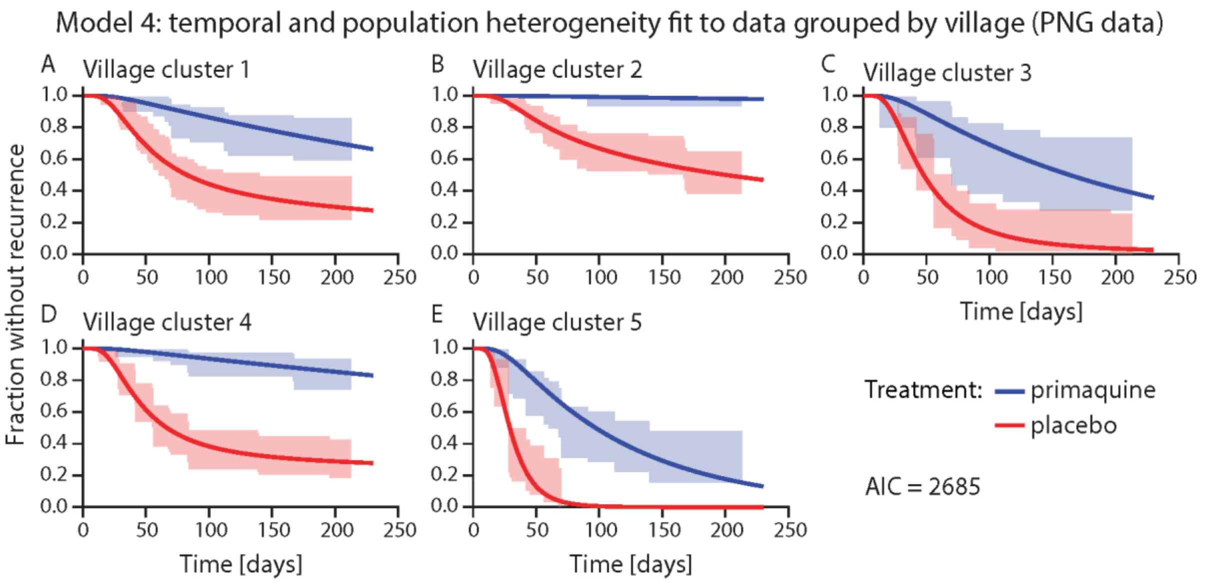


**Fig. S4** Fit of the temporal and population heterogeneity model to the first *P vivax* infection by PCR in the PNG data grouped by village. All villages were fit simultaneously with the same drug washout time distribution, the rate of new infections and relapses were allowed to vary between villages. The lines indicate the model fit and the shaded area the 95% confidence region from the data. For the parameters of the model fit see **Table S8**.

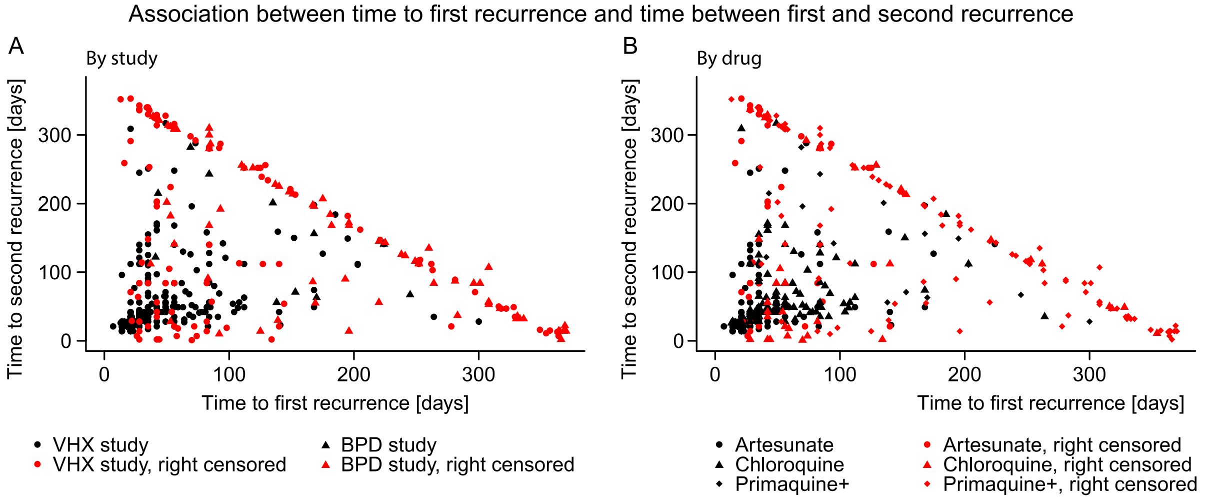


**Fig. S5** Association between time to first recurrence and time from first to second recurrence in the Thailand-Myanmar data. Different symbols represent different studies. **B** The symbols represent the different drugs. The Spearman correlation between time to first recurrence and time from first to second recurrence can be found in **Table S11**.

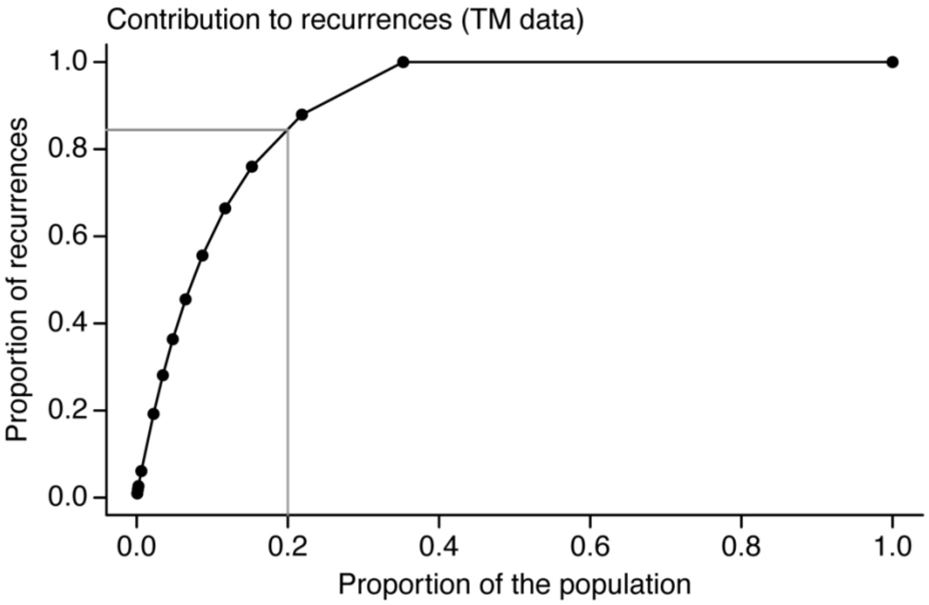


**Fig. S6** Contribution to recurrences for all patients in the Thailand-Myanmar data. This figure shows which percent of recurrences is caused by which percent of the population. Each dot represents the number of recurrences from 14 to 0 (from left to right), i.e., the first dot represents the percent of the population with at least 14 recurrences (x-axis) and the percent of recurrences caused by the patients with at least 14 recurrences (y-axis). The 20% of the population with the highest number of recurrences cause almost 85% of all recurrences (gray lines). This figure includes all patients, i.e., those who were treated for blood-stage infections only and those who were treated for both blood- and liver-stage infections. For the contribution to relapses for patients who were treated only for blood-stage infections see **Fig. 6**A.

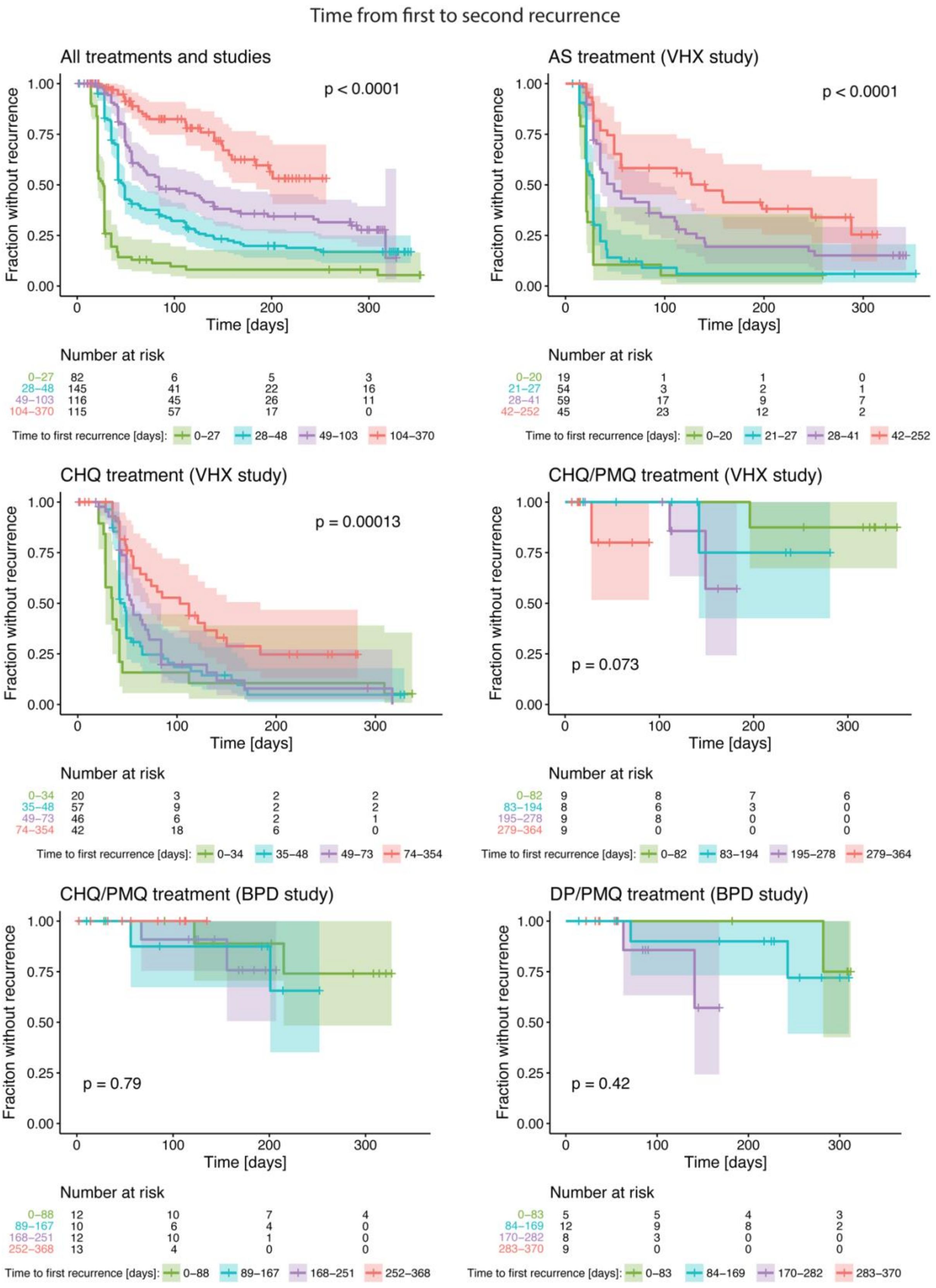


**Fig. S7** Time from first to second recurrence by the time to first recurrence quartiles for the different antimalarial treatments and studies in the Thailand-Myanmar data. The shaded areas are the 95% confidence regions for the survival curves and the p-value of the log-rank test for the comparison of the survival curves is shown in each panel.

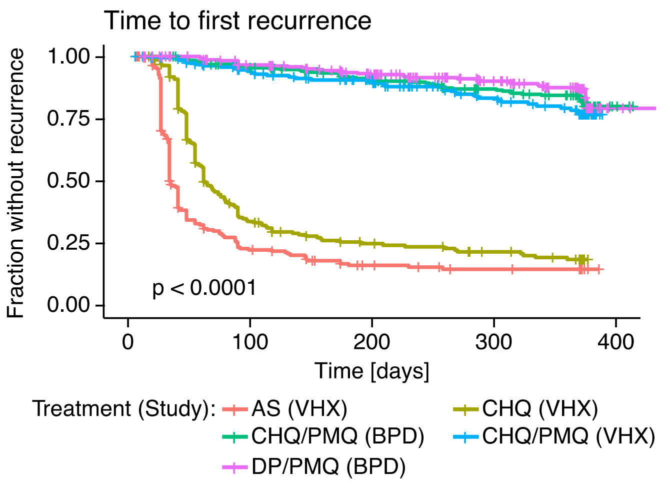


**Fig. S8** Time to first recurrence for the Thailand-Myanmar data grouped by antimalarial treatment and study. The p-value of the log-rank test for the comparison of all survival curves is shown in the lower left corner of the plot. The survival curves are significantly different, however, there is no significant difference between the survival curves of individuals treated with primaquine (CHQ/PMQ and DP/PMQ; p-value 0.074). The artesunate (AS) and chloroquine (CHQ) treatment survival curves are significantly different (p-value <0.0001). This difference is due to the different dynamics within the first 50 days as excluding recurrences and censoring within the first 50 days gives a non-significant p-value of 0.14.

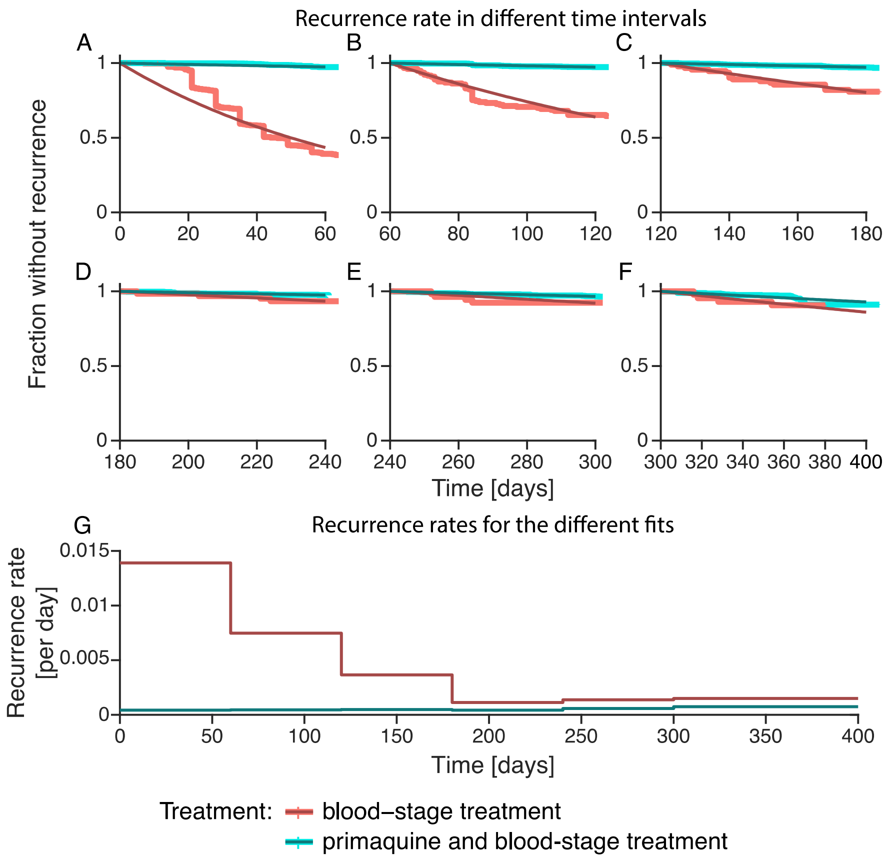


**Fig. S9** Recurrence rate on days 0-60, 60-120, 120-180. 180-240, 240-300, and 300-400 in the Thailand-Myanmar data. **A-F** Model fit to a part of the data grouped by blood-stage treatment and primaquine and blood-stage treatment. The data are shown in the lighter color, the model fits in a darker color. **G** Recurrence rates for the two different treatment groups and the fits shown in **A-F** over time. The estimated percentage of blood-stage infections that are relapses are 96.9%, 93.8%, 86.9%, 62.7%, 57.9%, and 50.8% for the fit to days 0-60, 60-120, 120-180. 180-240, 240-300, and 300-400, respectively.

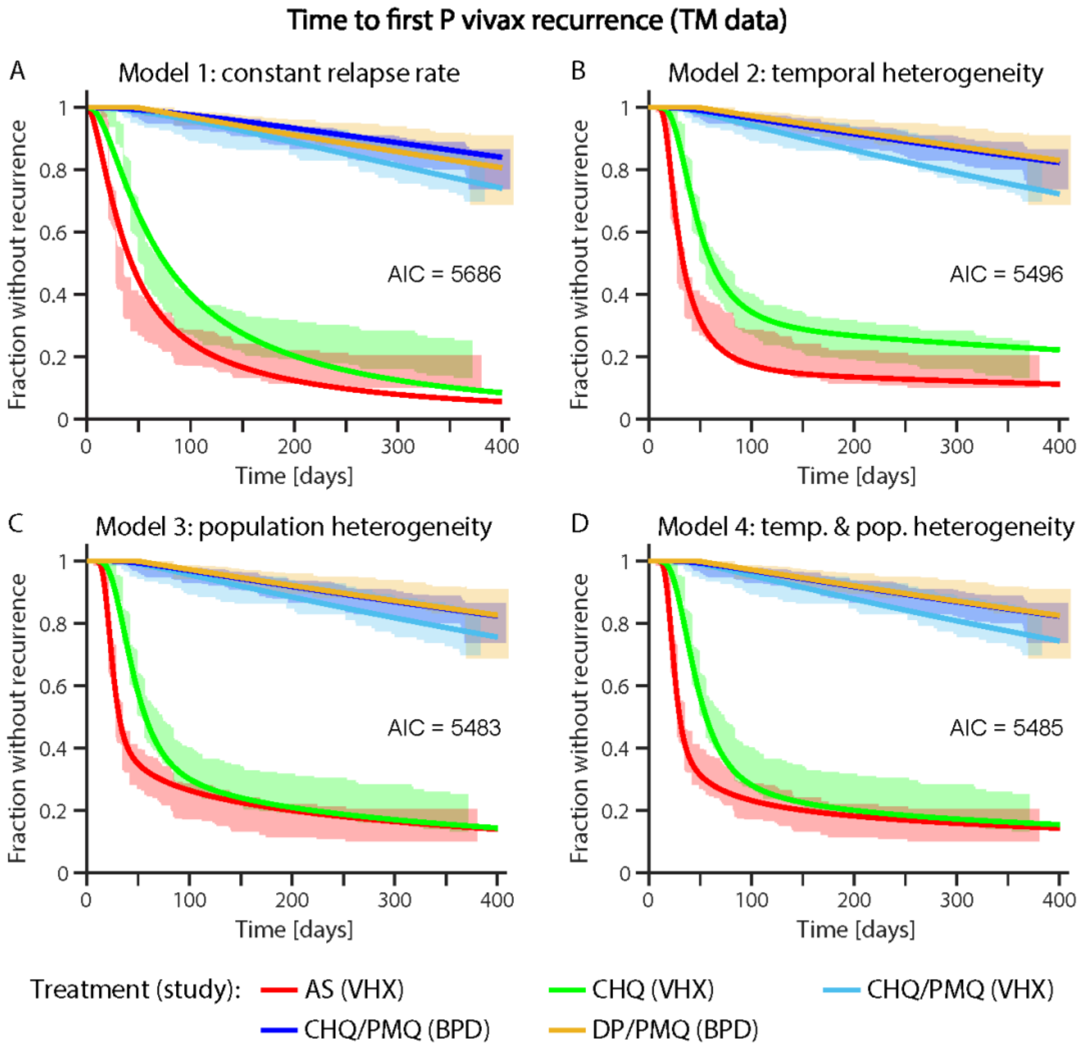


**Fig. S10** Model fit of the constant relapse rate, temporal heterogeneity, the population heterogeneity, and the temporal & population heterogeneity model to the first recurrence time in the Thailand-Myanmar data. The shaded areas are the 95% confidence regions from the data. Abbreviations: AS artesunate, CHQ chloroquine, CHQ/PMQ chloroquine and primaquine, DP/PMQ dihydroartemisinin-piperaquine and primaquine, VHX Vivax History study, BPD best Primaquine Dose study. For the parameters of the model fit see **Table S14**, **Table S15**, **Table S16**, and **Table S17**.

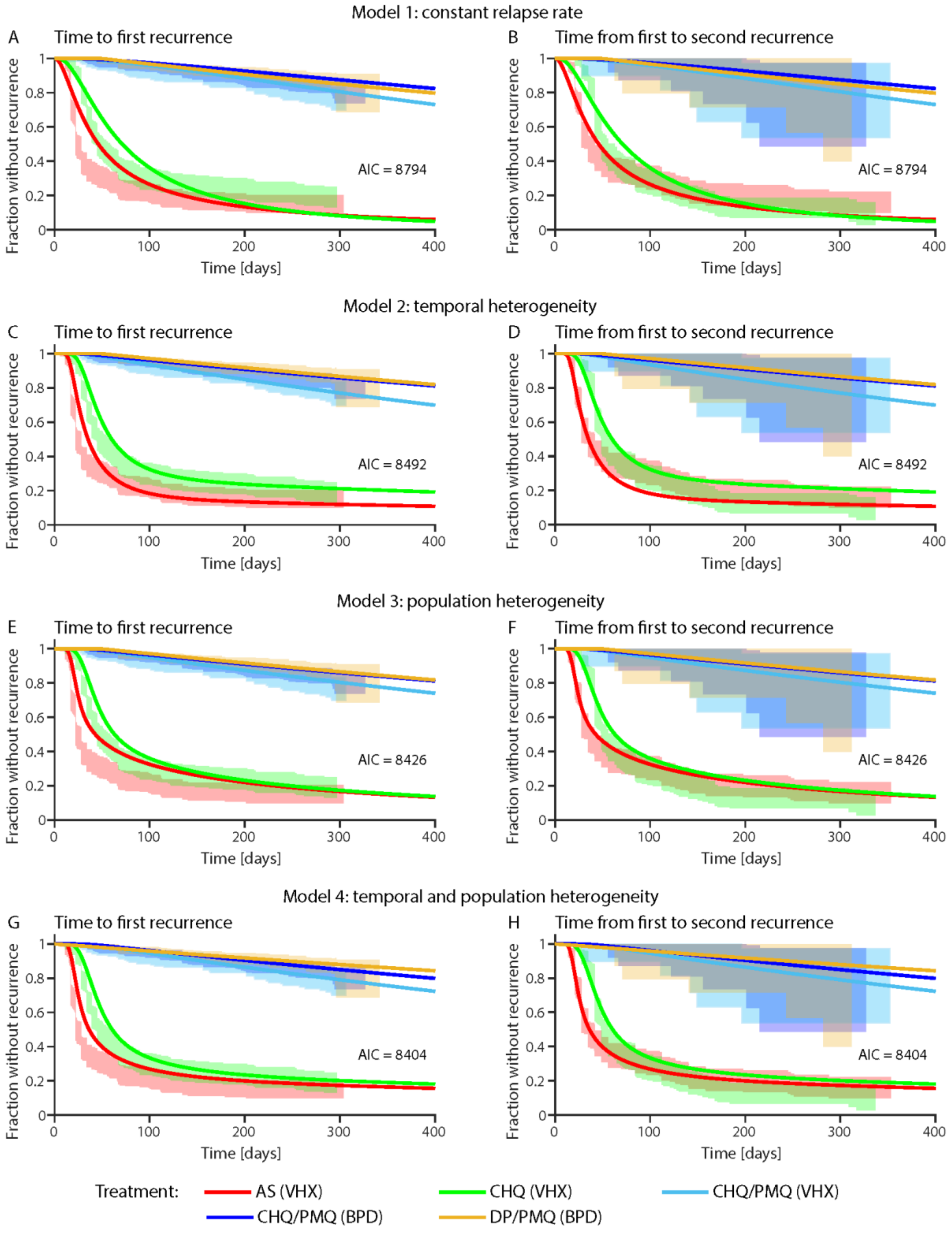


**Fig. S11** Model fit of the constant relapse rate, temporal heterogeneity, the population heterogeneity, and the temporal and population heterogeneity model to the first and second recurrence times in the Thailand-Myanmar data simultaneously. The shaded areas are the 95% confidence regions from the data. The comparison of models 3 and 4 with the likelihood-ratio test indicates that model 4 fits the data significantly better than model 3 (p-value < 0.0001). Abbreviations: AS artesunate, CHQ chloroquine, CHQ/PMQ chloroquine and primaquine, DP/PMQ dihydroartemisinin-piperaquine and primaquine, VHX Vivax History study, BPD best Primaquine Dose study. For the parameters of the model fit see **Table S18**, **Table S19**, **Table S20**, and **Table S21**.

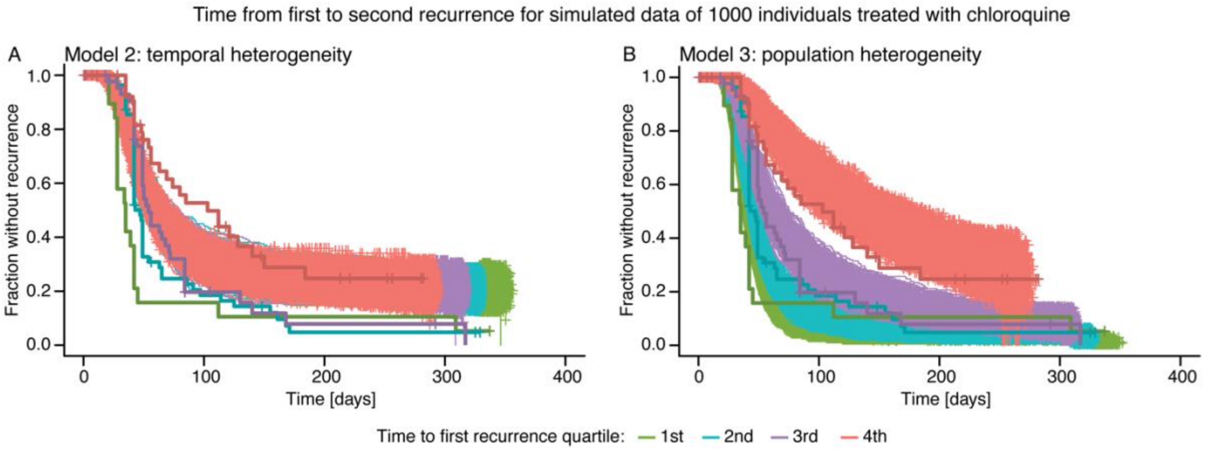


**Fig. S12** Comparison of simulated data for models 2 and 3 with the original data from the Thailand-Myanmar border region. This figure shows the survival curves of 1000 simulated populations of 1000 simulated individuals with chloroquine treatment (thin lines). The survival curves from the original data are shown for comparison as bold and darker lines. Both the simulated data and the original data are grouped by the time to first recurrence quartiles. The parameters used for this simulation are the parameter values of model fits to the TM data 1st and 2nd recurrence (see **Table S19** and **Table S20**). **A** Data simulated using the temporal heterogeneity model. **B** Data simulated using the population heterogeneity model.

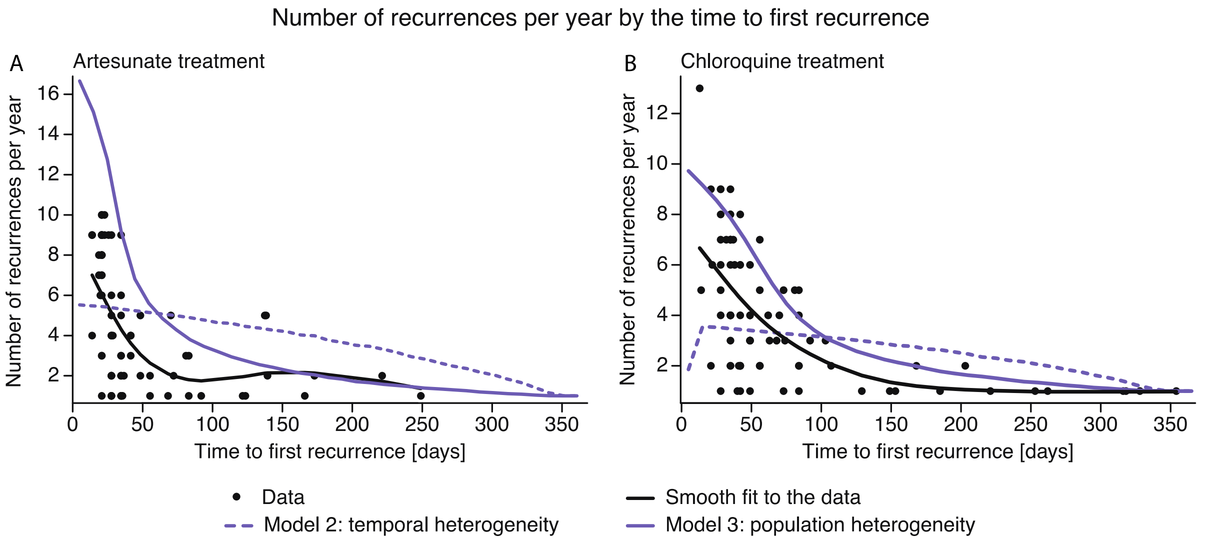


**Fig. S13** Number of recurrences per year in the Thailand-Myanmar data and simulated data for artesunate (**A**) and chloroquine (**B**) treatment. Each dot represents an individual from the data who had an overall follow-up time of at least 1 year. Patients with a follow-up less than one year were excluded. The smooth fit to the data is a smooth spline (fit using the "smooth.spline" function in R with 4 degrees of freedom). The number of recurrences per year for the models were calculated from the simulated data as the average number of recurrences over all individuals with time to first recurrence within the same 10-day time interval.

### Additional methods

#### Models

We constructed mathematical models for *P vivax* recurrences. For the scheme of the models see **Fig. S14**. Individuals are protected due to the prophylactic effect of antimalarial treatment at enrolment. The drug washout time is lognormally distributed and after drug washout individuals are susceptible to both new, mosquito-borne infections and relapses. All models include a constant infection rate but differ in how relapses are modelled (see also the Methods section in the main text).

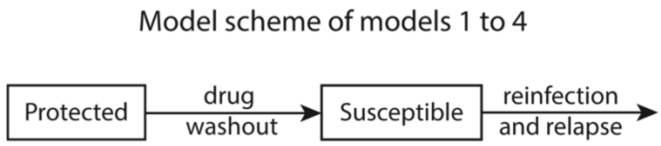


**Fig. S14** Model scheme of models 1 to 4. At enrolment patients are treated and thus protected due to the prophylactic effect of the antimalarials. After drug washout, patients are susceptible to both new infections and relapses. Models 1 to 4 all include a lognormal distributed drug washout time and a constant infection rate. The models differ in their relapse rate.

##### Model 1: constant relapse rate

The relapse rate is constant and the same for all individuals. Thus, the time to the next relapse is exponentially distributed and the fraction of susceptible individuals $S(t)$ at time $t$ is given by the following ODE:

$$\frac{d}{dt}\,S(t) = w(t;\mu,\sigma) - (r+n)\,S(t), \qquad S(0)=0,$$

where $w(t;\mu,\sigma)$ is the probability density function of the lognormal distribution with parameters $\mu$ and $\sigma$, $r$ is the constant relapse rate, and $n$ is the constant infection rate. This model equation describes that individuals become susceptible after the lognormally distributed drug washout time ($w(t;\mu,\sigma)$) and leave the compartment of susceptible individuals after a relapse occurring at rate $r$ or a new infection occurring at rate $n$. Initially, all individuals are protected, thus $S(0) = 0$.

##### Model 2: temporal heterogeneity

The relapse rate in model 2 is a time-dependent relapse rate given by $r(t) = Ie^{-dt}$ (see main text for more details). The model equation is similar to the model equation for model 1:

$$\frac{d}{dt}\,S(t) = w(t;\mu,\sigma) - (r(t)+n)\,S(t), \qquad S(0)=0,$$

where $w(t;\mu,\sigma)$ is the probability density function of the lognormal distribution with parameters $\mu$ and $\sigma$, $r(t) = Ie^{-dt}$ is the time-dependent relapse rate, and $n$ is the constant infection rate.

##### Model 3: population heterogeneity

Model 3 takes population heterogeneity in relapses into account as a distribution in relapse rates. Each individual has a random relapse rate drawn from a lognormal distribution. In order to simplify the numerical solution of model 3, we group the population into 'relapse risk groups' of equal size (see **Fig. S15**). As we use percentiles of the relapse risk distribution to define the relapse risk groups, all relapse risk groups have the same size (meaning the same proportion of the population is in each of the relapse risk groups). The relapse rate of each risk group is the median relapse rate of this group. Thus, for $k$ relapse risk groups with relapse rates $r_i$ the model equation for risk group $i$ is given by:

$$\frac{d}{dt} S_i(t) = w(t;\mu,\sigma)/k - (r_i + n)\, S_i(t), \qquad S_i(0) = 0,$$

where $S_i(t)$ is the fraction of susceptibles who are in risk group $i$ ($i \in \{1,2,\dots,k\}$) at time $t$, $k$ is the number of relapse risk groups, $r_i$ is the median relapse rate of group $i$, and $n$ is the constant infection rate. This model equation describes that individuals are equally distributed to the risk groups, thus $w(t;\mu,\sigma)/k$ is the fraction of individuals who are susceptible and in risk group $i$ after the lognormal distributed drug washout time. Individuals leave the compartment of susceptible individuals after a relapse (at rate $r_i$) or a new infection (at rate $n$). The overall fraction of susceptible individuals at time $t$, $S(t)$, is then given by the sum of all susceptible individuals in the different risk groups:

$$S(t) = \sum_{i=1}^{k} S_i(t).$$

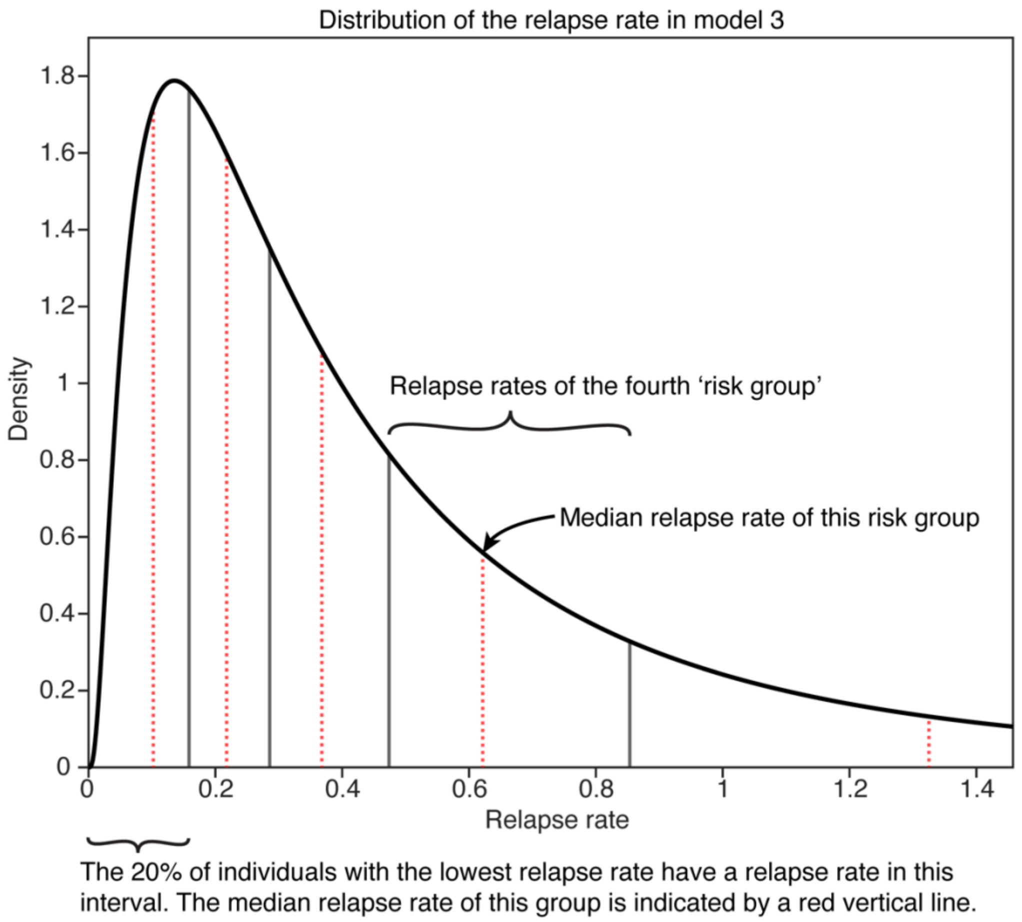


**Fig. S15** Scheme of the distribution of the relapse rates in model 3. Relapse rates in model 3 are lognormally distributed (black curve). The population is divided into 'relapse risk groups' of equal size (vertical grey lines). In this scheme there are five relapse risk groups. When numerically solving the model equation, individuals from the same relapse risk group are considered to have the same relapse rate which is chosen as the median relapse rate of that group (vertical red dotted lines).

**Model 4: temporal and population heterogeneity**

Model 4 takes both population heterogeneity and temporal heterogeneity in relapses into account as a combination and extension of models 2 and 3. As for model 3, we group the population in $k$ different relapse risk groups of equal size. We use again percentiles of the relapse risk distribution (a lognormal distribution) to define the relapse risk groups. Individuals in the same relapse risk group have the same initial relapse risk that decreases over time as in model 2. Thus, the relapse risk of group $i$ is given by:

$$r_i(t) = I_i e^{-dt},$$

where $I_i$ is the initial relapse risk for relapse risk group $i$ and $d$ is the relapse risk decay rate that we assume to be the same for all individuals regardless of their initial relapse risk. Model 4 is given by:

$$\frac{d}{dt} S_i(t) = w(t; \mu, \sigma)/k - (r_i(t) + n)\, S_i(t), \qquad S_i(0) = 0,$$

where $S_i(t)$ is the fraction of susceptible individuals that are in risk group $i$ ($i \in \{1,2,\ldots,k\}$) at time $t$, $k$ is the number of relapse risk groups, $r_i(t) = I_i e^{-dt}$ is the time-dependent relapse rate of group $i$, and $n$ is the constant reinfection rate. As for model 3, the overall fraction of susceptible individuals at time $t$, $S(t)$, is the sum of all susceptible individuals in the different risk groups.

#### Models for two recurrences

To fit the models not only to the first recurrence after enrolment but to the first and second recurrence (only for the Thailand-Myanmar data because the Papua New Guinea data does not contain multiple recurrence times), we extend the models to take two recurrences into account. The model scheme is shown in **Fig. S16**.

For models 1 to 4, we extend the models with an additional compartment for protected and susceptible individuals (see **Fig. S16**). We make the following model assumptions:

- After the first recurrence, individuals are again protected. Thus, it is assumed that all recurrences are detected and immediately treated. We do not explicitly consider blood-stage infections and their duration.
- The drug washout time and time to recurrence follow the same distribution for the first and second recurrence. Thus, there is either no significant change in the hypnozoite number or the relapse rate is independent of the hypnozoite number. There is also no seasonality or changing of the infection rate over time.

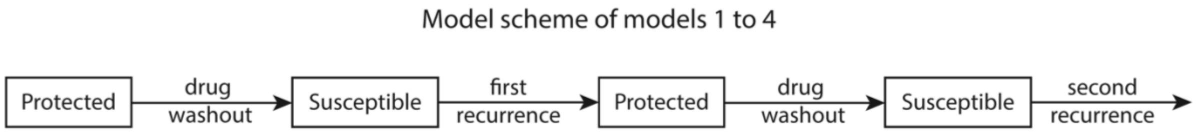


**Fig. S16** Model scheme of models 1 to 4 for two recurrences. After the first recurrence, we assume that all individuals are treated with the same drug as at enrolment. Both the drug washout rate and the recurrence rate for the second recurrence are the same as for the first recurrence for models 1 to 4.

**Parameters of the models**

The Thailand-Myanmar data consists of two different studies and individuals treated with different antimalarials. Different parameters of the models may vary by study or antimalarial treatment (see **Table S22**).

For the constant infection rate, we considered both the case that the infection rate is the same for the two different studies and the case that is different (see **Table S23**).

The drug washout time depends on the antimalarial treatment. In the case that individuals received a treatment with a combination of different drugs, the drug with the longer half-life determines the drug washout time. The half-lifes of the different antimalarials are 20-45 minutes for artesunate (AS) [1], 4-6 hours for primaquine (PMQ) [2, 3], approx. 3 weeks for dihydroartemisinin-piperaquine (DP) [4, 5], and 1-2 months for chloroquine (CHQ) [6]. Thus, for individuals treated with chloroquine and primaquine (CHQ/PMQ) the drug washout time is determined by chloroquine. For individuals treated with dihydroartemisinin-piperaquine and primaquine (DP/PMQ) the drug washout time is determined by dihydroartemisinin-piperaquine. Note that recurring *P vivax* infections were treated with the same antimalarial as at enrollment in the VHX study and with the standard chloroquine and primaquine in the BPD study. Thus, all individuals except the group treated DP/PMQ in the BPD study were treated with the same antimalarial at each recurrence. For simplicity, we fitted a separate drug washout time distribution for the DP/PMQ group, thus we assume that these individuals are treated with the same antimalarial at each treatment or at least an antimalarial with the same washout time distribution.

For the relapse parameters, we distinguish between individuals who received blood-stage treatment only (AS and CHQ) and those who also received primaquine (CHQ/PMQ and DP/PMQ). Since primaquine is a radical cure killing parasites of all stages including hypnozoites [7], individuals who were treated with primaquine are expected to not have any relapses.

**Parameter dependency groups in the model fits to the Thailand-Myanmar data**

| Treatment | Study (group for new infection rate) | Drug washout distribution group | Relapse group |
|---|---|---|---|
| Artesunate | VHX | AS | Blood-stage treatment |
| Chloroquine | VHX | CHQ | Blood-stage treatment |
| Chloroquine & Primaquine | VHX | CHQ | - |
| Chloroquine & Primaquine | BPD | CHQ | - |
| Dihydroartemisinin-Piperaquine & Primaquine | BPD | DP | - |

**Table S22** This table shows how the parameters for new infections, the drug washout distribution, and relapses depend on treatment and study in the Thailand-Myanmar data. The parameters are the same if they are in the same group, e.g., all individuals treated with chloroquine have the same drug washout time distribution regardless of whether they were treated with chloroquine and primaquine or only with chloroquine.

The parameters for each model and each data set are given below.

**Model 1: constant relapse rate**

**PNG data:** Since in this model both the infection rate and the relapse rate are constant, we sum them to a constant recurrence rate (each individually would not be identifiable). The model contains two parameters for the drug washout time distribution, the recurrence rate for patients treated for blood-stage infection only, and the recurrence rate for patients treated for both blood- and liver-stage infection.

**PNG data by village**: The model contains two parameters for the drug washout time distribution for all villages and the recurrence rates for patients treated for blood-stage infection only and patients treated for both blood- and liver-stage infections for each of the 5 villages. Overall, model 1 fit to the PNG data by village contains 12 parameters.

**Thailand-Myanmar data** (for the case of two different infection rates): The model contains six parameters for the drug washout time, the mean and standard deviation for the lognormal distribution of drug washout times for AS, CHQ, and DP, respectively. Thus, we have the recurrence rate for individuals in the VHX study who received blood-stage treatment, the recurrence rate for the VHX study with primaquine treatment, and the recurrence rate for the BPD study with primaquine treatment. Overall, this model has 9 parameters.

**Model 2: temporal heterogeneity**

**PNG data:** The model contains two parameters for the drug washout time distribution, the rate of new infections, and for the time-dependent relapse rate $r(t) = Ie^{-dt}$, the initial relapse rate ($I$) and the rate of decay of the relapse rate ($d$) for blood-stage treatment. Overall, the model contains 5 parameters.

**PNG data by village**: The model contains two parameters for the drug washout time distribution for all villages and the rate of new infections, the initial relapse rate, and the exponential decay rate of the relapse rate for each of the 5 villages. Overall, model 2 fit to the PNG data by village contains 17 parameters.

**Thailand-Myanmar data** (for the case of two different infection rates): The model contains the mean and standard deviation for the lognormal distribution of drug washout times for AS, CHQ, and DP, respectively, and two infection rates for VHX and BPD. For the time-dependent relapse rate $r(t) = Ie^{-dt}$, we have the initial relapse rate ($I$) and the rate of decay of the relapse rate ($d$) for blood-stage treatment. Overall, there are 10 parameters in this model.

**Model 3: population heterogeneity**

**PNG data:** The model contains two parameters for the drug washout time distribution, the rate of new infections, and two parameters for the distribution of relapse rates.

**PNG data by village**: The model contains two parameters for the drug washout time distribution for all villages and the rate of new infections and two parameters for distribution of relapse rates for each of the 5 villages. Overall, model 3 fit to the PNG data by village contains 17 parameters.

**Thailand-Myanmar data** (for the case of two different infection rates): The model contains the mean and standard deviation for the lognormal distribution of drug washout times for AS, CHQ, and DP, respectively, and two infection rates for VHX and BPD. The relapse rate is lognormal distributed. Thus, the model also contains the mean and standard deviation for the relapse rate distribution for blood-stage. Overall, model 3 has 10 parameters.

This models also contains a parameter that is integer-valued, the number of relapse risk groups. We fit the model for different numbers of relapse risk groups and compared the model fits (see **Fig. S21**).

**Model 4: temporal and population heterogeneity**
**PNG data:** The model contains two parameters for the drug washout time distribution, the rate of new infections, two parameters for the distribution of the initial relapse rates, and the exponential decay rate of the relapse rate.
**PNG data by village**: The model contains two parameters for the drug washout time distribution for all villages and the rate of new infections, two parameters for distribution of relapse rates, and the decay rate of the relapse rate for each of the 5 villages. Overall, model 4 fit to the PNG data by village contains 22 parameters.
**Thailand-Myanmar data** (for the case of two different infection rates): The model contains the mean and standard deviation for the lognormal distribution of drug washout times for AS, CHQ, and DP, respectively, and two infection rates for VHX and BPD. The initial relapse rate is lognormal distributed and decays exponentially. Overall, model 4 has 11 parameters.
As model 3, this models also contains a parameter that is integer-valued, the number of relapse risk groups.

For a list of all the parameter values as well as their maximum likelihood estimates and their 95% confidence intervals, see **Table S1** to **Table S4** for the PNG data, **Table S5** to **Table S8** for the PNG data by village, and **Table S18** to **Table S21** for the Thailand-Myanmar data.

**Likelihood function for fitting to the first and second recurrence time simultaneously (Thailand-Myanmar data only)**

We use the following notation for the likelihood function:

- $p$: vector of parameters for the model
- $D$: data, $D_i$: data for individual $i$ (there are overall $N$ individuals who can be divided into $N_0$ individuals with no recurrences, $N_1$ individuals with 1 recurrence, and $N_2$ individuals with at least 2 recurrences)
- $j$: relapse risk group number out of different $r$ risk groups of equal size (**Fig. S15**)
- $R_i$: relapse risk group of individual $i$
- $U_j(t)$: probability that an individual in risk group $j$ has a recurrence more than $t$ days after the previous recurrence (i.e., stays uninfected for at least $t$ days).
  Note that for models 1 to 4 there is no difference between $U_j(t)$ for the first and the second recurrence as the drug washout time, relapses, and recurrences are assumed to have the save distribution for the first and the second recurrence.
- $G_j(t) = U_j(t-\Delta) - U_j(t)$: probability that an individual in risk group $j$ has a recurrence between day $t-\Delta$ (the day of the last follow-up before a recurrence, $\Delta$ depends on the follow-up scheme) and day $t$ (follow-up visit with a recurrence) after the previous recurrence.

The likelihood function is given by:

$$L(p|D) = P(D|p) = \prod_{i=1}^{N} P(D_i|p) = \prod_{i=1}^{N}\left[\sum_{j=1}^{r} P(D_i|R_i = j) \times P(R_i = j)\right],$$

where we assume that individuals are independent, we split up the population into the different 'relapse risk' groups $R_i$, and in the last step (and in the following) we omit the parameters $p$ to keep the notation simpler.

For each individual, the data $D_i$ contains the number of recurrences, recurrence times, and times of censoring. Thus, each individual will fall into one of the three groups below (omitting the index $i$ for individual $i$ in our notation for simplicity):

- 0 recurrences: denoted as $n = 0$, where $n$ is the number of recurrences.
- 1 recurrence at time $t_1$: denoted as $(n = 1) \cap t_1$.
- At least 2 recurrences at times $t_1$ and $t_2$ (where $t_1$ is the time from the beginning of the study to the first recurrence and $t_2$ is the time from the first recurrence to the second recurrence): denoted as $(n \geq 2) \cap t_1 \cap t_2$

Next, we determine $P(D_i|R_i = j)$ for each of these three cases:

- 0 recurrences case:
  $$P(n = 0|\, R = j) = U_j(T),$$
  where $T$ is the overall follow-up time.
- 1 recurrence case:
  $$\begin{aligned} P((n = 1) \cap t_1|\, R = j) &= P(n = 1 \mid t_1, R = j) \times P(t_1|R = \, j) \\ &= U_j(T - t_1) \times G_j(t_1). \end{aligned}$$
- At least 2 recurrences case:
  $$\begin{aligned} P((n \geq 2)\ \cap\ t_1 \cap\ t_2 \mid j) &= P(n \geq 2|\, t_1, t_2, j) \times P(t_1 \cap t_2|\, j) \\ &= G_j(t_1) \times G_j(t_2 - t_1), \end{aligned}$$

where in the last step $P(n \geq 2|\, t_1, t_2, j) = 1$ as the probability to have at least two recurrences given the time of two recurrences is 1. Alternatively, this can also be derived mathematically in the following way:

$$P(n \geq 2|\, t_1, t_2, R = j) = P(n = 2|\, t_1, t_2, R = j) + P(n > 2|\, t_1, t_2, R = j)$$
$$= U_j(T - (t_1 + t_2)) + \left[1 - U_j\big(T - (t_1 + t_2)\big)\right] = 1$$

as the probability to have exactly two recurrences is equal to the probability to have no more recurrences in the remaining time (from the second recurrence to censoring) and the probability to have more than two recurrences is the probability to have at least one recurrence in the remaining time (i.e., to not have no recurrences).

Now, we know $P(D_i|R_i = j)$ for each of the three cases. Splitting up the population into individuals with 0, 1, or at least 2 recurrences and using the above formulas for the likelihood function and $P(D_i|R_i = j)$, we obtain the following likelihood function:

$$L(p|D) = \prod_{i=1}^{N_0}\left[\sum_{j=1}^{r} U_j(T_i) \times P(R_i = j)\right] \times \prod_{i=1}^{N_1}\left[\sum_{j=1}^{r} G_j\big(t_{i,1}\big) \times U_j(T - t_1) \times P(R_i = j)\right]$$
$$\times \prod_{i=1}^{N_2}\left[\sum_{j=1}^{r} G_j(t_{i,1}) \times G_j(t_{i,2}) \times P(R_i = j)\right].$$

The general loglikelihood function is then:

$$l(p|D) = \sum_{i=1}^{N_0} \log\left[\sum_{j=1}^{r} U_j(T_i) \times P(R_i = j)\right]$$
$$+ \sum_{i=1}^{N_1} \log\left[\sum_{j=1}^{r} G_j\big(t_{i,1}\big) \times U_j(T_i - t_{i,1}) \times P(R_i = j)\right]$$
$$+ \sum_{i=1}^{N_2} \log\left[\sum_{j=1}^{r} G_j\big(t_{i,1}\big) \times G_j(t_{i,2}) \times P(R_i = j)\right].$$

In the case of only one risk group for the entire population, i.e., for models 1 and 2, the loglikelihood function simplifies to:

$$l(p|D) = \sum_{i=1}^{N_0} \log[U(T_i)] + \sum_{i=1}^{N_1} \log\big[U\big(t_{i,1}\big) \times U(T_i - t_{i,1})\big] + \sum_{i=1}^{N_2} \log\big[G\big(t_{i,1}\big) \times G(t_{i,2})\big].$$

In the case that individuals are equally distributed to the different risk groups (by using percentiles of the relapse rate distribution), i.e., in models 3 and 4, we have the following simplified loglikelihood function:

$$l(p|D) = \sum_{i=1}^{N_0} \log\left[\sum_{j=1}^{r} U_j(T_i)\right] + \sum_{i=1}^{N_1} \log\left[\sum_{j=1}^{r} G_j(t_{i,1}) \times U_j(T_i - t_{i,1})\right] + \sum_{i=1}^{N_2} \log\left[\sum_{j=1}^{r} G_j(t_{i,1}) \times G_j(t_{i,2})\right] + (N_0 + N_1 + N_2) \times \log\left[\frac{1}{r}\right].$$

The last term in the loglikelihood function for model 3 comes from $P(R_i = j)$ as it holds that

$$P(R_i = j) = \frac{1}{r}$$

for all individuals $i$ and all risk groups $j$. In models 3 and 4, the risk groups were chosen by the percentiles of the relapse rate distribution, i.e., a fraction $1/r$ of the population is in each of the risk groups.

**Model fits**

In order to fit the models to the time-to-recurrence data for the first (and to the second for the Thailand-Myanmar data) recurrence, we numerically solve the model equations (see main text) using the ODE solver ode15s in Matlab (version R2018b) [8].
We obtain the fraction of susceptible individuals at time $t$, $S(t)$, as the numerical solution of the model equation. The fraction of individuals that remain uninfected at time $t$ is then given as all the individuals who are still protected by the antimalarial treatment and all the susceptible individuals who have not yet been reinfected or had a relapse, i.e.,

$$U(t) = \left(1 - \int_0^t w(\tau; \mu, \sigma)\, d\tau\right) + S(t),$$

where $w(t; \mu, \sigma)$ is the probability density function of the lognormal distribution of drug washout times with parameters $\mu$ and $\sigma$.
We interpret $U(t)$ as the probability to be uninfected until time $t$ and use it to define the probability to have an infection at the visit on day $t$ ($G(t)$) as in the main text:

$$G(t) = U(t - \Delta) - U(t),$$

where $t - \Delta$ is the time of the last follow-up visit before day $t$.
Both $U(t)$ and $G(t)$ depend on the model parameters, $G(t)$ also depends on the follow-up scheme, and for the population heterogeneity model they both depend on the number of relapse risk groups. We tried either one rate of new infections or two different rates of new infections for the two different studies in the Thailand-Myanmar data, different follow-up schemes and different numbers of relapse risk groups in the population heterogeneity model (see Supplementary results). In the end, we chose a daily follow-up scheme, 10 different relapse risk groups, two rates of new infections in the Thailand-Myanmar data, and one rate of new infections for the PNG data for all model comparisons and all data sets.
With $U(t)$ and $G(t)$ we can use the above loglikelihood function to fit our models to the first and second recurrence time in the Thailand-Myanmar data and obtain Maximum Likelihood Estimates (MLEs) for the parameter values. We do so by selecting random initial parameter values and minimizing the negative loglikelihood function using the Matlab function fmincon. In order to assure that we obtain a good fit, we minimize the negative loglikelihood function for 100 random initial parameter vectors and the MLE of the parameters for the fit to the first recurrence only (see **Table S14**, **Table S15**, **Table S16**, and **Table S17**).
We fit to the first recurrence times in the same way as to the first and second recurrence time in the Thailand-Myanmar data. However, instead of the above loglikelihood function, we used the following simpler loglikelihood function:

$$l(p|D) = \sum_{i=1}^{N_1} \log[U(t_i - \Delta) - U(t_i)] + \sum_{i=1}^{N_0} \log\,[U(T_i)],$$

where $N_0$ and $N_1$ are the numbers of individuals with zero and at least one recurrence, respectively, $U(t)$ is the probability to be uninfected until time $t$, $t_i$ is the time of the first recurrence of individual $i$, $t_i - \Delta$ is the time of the last follow-up visit before day $t_i$, and $T_i$ is the follow-up time of individual $i$. As for the model comparison for the fit to the first and second recurrence time, we use daily follow-up and 10 relapse risk groups in the population heterogeneity model.
We compare the model fits using the Akaike Information Criterion (AIC) that is given by

$$AIC = 2 \times \left(-l(p|D)\right) + 2 \times n_{par},$$

where $-l(p|D)$ denotes the negative loglikelihood and $n_{par}$ denotes the number of parameters.
We also compared model fits of model 4 with model 3 using the likelihood-ratio test and found that model 4 is a significantly better model (p-value < 0.0001).

**Confidence Intervals**

We computed confidence intervals using bootstrapping and the percentile method. We drew individuals from the data with replacement and fitted each model to the new data as described above. However, we used only 10 random initial parameter values and the best fitting parameter values for the original data for fitting each model to the bootstrapped data (for efficiency and time reasons). This was repeated 1000 times. The 95% confidence interval for each parameter is the 95th percentile of the MLE of the best fitting parameters for the bootstrapped data.

**Model simulations**

We simulated 1,000 cohorts of 1,000 individuals for 1 year and artesunate or chloroquine using models 1 to 3. The parameters for the model simulation are the MLEs of the parameters from the fit to the first and second recurrence time in the Thailand-Myanmar data (see **Table S18**, **Table S19**, and **Table S20**). The detailed description of the model simulation method for each model is given below.

**Model 1: constant relapse rate**
For each individual a drug washout time is drawn from a lognormal distribution with the appropriate parameters depending on the antimalarial treatment. Since the recurrence rate is constant, the time from drug washout to a recurrence is exponentially distributed. Thus, the recurrence time is drawn from an exponential distribution with mean 1/recurrence rate. The time of the first recurrence is then the sum of the drug washout time and the time from drug washout to recurrence. This process is repeated until the individual has been simulated for 1 year.

**Model 2: temporal heterogeneity**
The drug washout time is drawn from a lognormal distribution as for model 1. In this model, the rate of new infections is constant, but the relapse rate is non-constant and decreases in time. For an event occurring at rate $r(t)$ the cumulative distribution function of the time to next event distribution is given by:

$$f(t) = 1 - e^{-\int_0^t r(x)dx}.$$

The recurrence rate is the sum of the rate of new infections and the relapse rate, thus the cumulative distribution function of the time to the next recurrence after drug washout is given by:

$$f(t) = 1 - e^{-\int_0^t n + Ie^{-dx}\, dx}.$$

Due to the prophylactic effect of the antimalarial treatment, there are no recurrences before drug washout. Thus, we shift the recurrence rate by the drug washout time $w$ and to take into account the blocking of relapses and new infections before drug washout. We obtain the following cumulative distribution function for the time to the next recurrence:

$$g(t,w) = 1 - e^{-\int_0^t n + Ie^{-d(x+w)}\, dx} = 1 - e^{-nt - \frac{I}{d}e^{-dw}\left(1-e^{-dt}\right)},$$

where $w$ is drug washout time.
Next, we simulate the time to the next recurrence using inverse transform sampling, i.e., we sample a number $x$ from a uniform distribution between 0 and 1 and estimate $g^{-1}(x,w)$ which is a sample of a random variable with cumulative distribution function $g(\cdot,w)$. We estimate $g^{-1}(x,w)$ by computing $g(\cdot,w)$ for a range of time points and choosing the time point $t$ for which $g(t,w)$ is closest to $x$. Thus, the time to the first recurrence is given by the estimate for $g^{-1}(x,w)$. As for model 1, this process is repeated until the individual has been simulated for 1 year.

**Model 3: population heterogeneity**
Each individual has a different relapse rate that is sampled from the lognormal distribution of relapse rates. The relapse rate is constant and each individual keeps the same relapse rate for

the entire simulated year. The drug washout time was computed as for models 1 and 2. Since both the rate of new infections and relapse rate are constant, the time to the next recurrence can be simulated as for model 1 by sampling from an exponential distribution.

Instead of simulating the time to the next recurrence, we can also simulate the time to the next new infection and the time to the next relapse in the same way as described above. The time to the next recurrence is then the smaller of these. This approach has the advantage that we know for each simulated recurrence whether it is a relapse or a new infection.
For model 1, we interpret the recurrence rate of primaquine treated patients as the rate of new infections. The rate of relapses is then the recurrence rate for artesunate or chloroquine treated patients minus the rate of new infections.
The time to relapse in model 2 can be simulated exactly as described above and the time to a new infection is a sample from an exponential distribution as the rate of new infections is constant.

### Additional results

##### Different infection rate for the different studies or one infection rate for both studies in the Thailand-Myanmar data

We fit the models to the data both using one infection rate (for new, mosquito-borne infections) as well as two different infection rates for the two different studies in the Thailand-Myanmar data (as Taylor et al. [9]). With two infection rates each model contains one more parameter and the AIC decreases by 5, 7, and 2 for models 1, 2, and 3, respectively (see **Table S23**). The model fits to first and second recurrence show very little difference for the fit with 1 or 2 infection rates (see **Fig. S17**). For this reason and due to the lower AICs, we do all model fits with two infection rates.

**Comparison of the model fit with 1 or 2 infection rates (Thailand-Myanmar data)**

| Model | AIC with 1 infection rate | AIC with 2 infection rates |
|---|---|---|
| 1: constant relapse rate | 8799 | 8794 |
| 2: temporal heterogeneity | 8499 | 8492 |
| 3: population heterogeneity | 8428 | 8426 |

**Table S23** Comparison of the model fit with 1 or 2 infection rates by AIC for the Thailand-Myanmar data. Including a second infection rate is one additional parameter for each model and improves the AIC by 5, 7, and 2 for models 1, 2, and 3, respectively. These model fits were done with daily follow-up and 10 relapse risk groups in model 3.

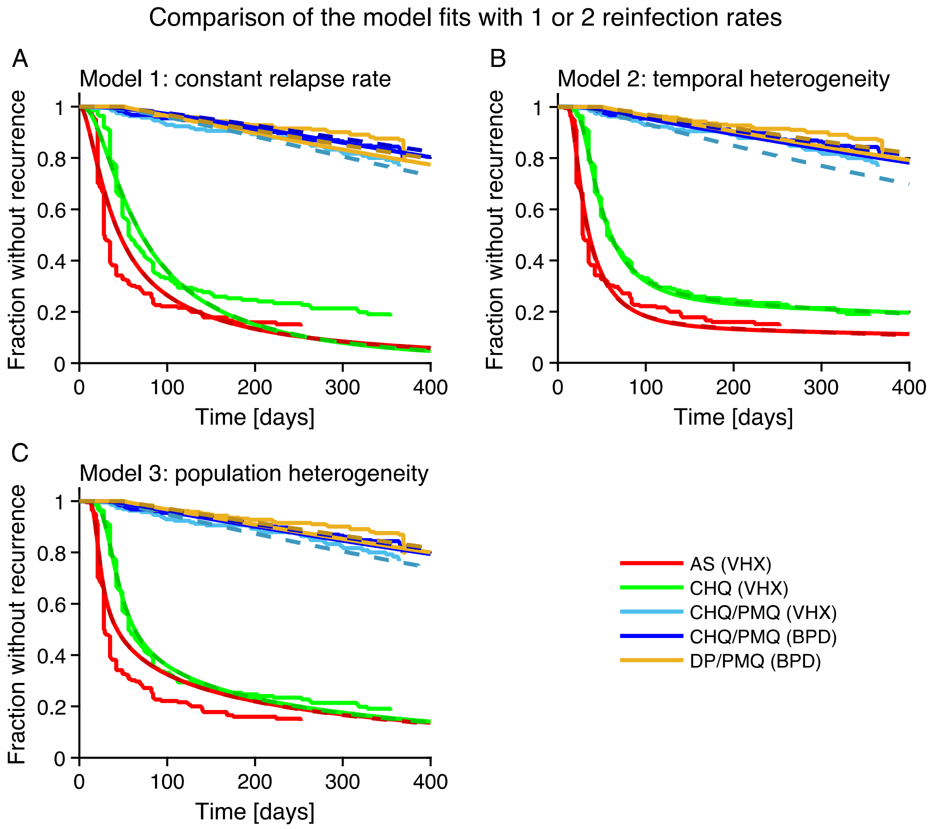


**Fig. S17** Comparison of the model fits with 1 infection rate for both studies and different infection rates for the two studies in the Thailand-Myanmar data. The data shown here are the time to the first recurrence survival curves for different antimalarial treatments. The model fit for one infection rate is shown in the same color as the corresponding data. Note that for one infection rate there is no difference between individuals treated with chloroquine and primaquine in the VHX study and the BPD study. The model fit for two different infection rates is shown as a dashed and darker line.

**Follow-up schemes in the Thailand-Myanmar data**

To compute the loglikelihood function and fit the models to the data, we need the follow-up scheme. According to Chu et al. [10], in the BPD study follow-up visits took place in weeks 2 and 4, then every four weeks. However, the data for the BPD study do not fit this follow-up scheme as recurrences were found also in weeks without follow-up visits. For the VHX study, it is not clear which follow-up scheme was used (the data also do not fit the follow-up scheme described for the BPD study). For this reason, we considered seven different follow-up schemes, fit the models using each of these follow-up schemes, and compare the model fits. We considered the following follow-up schemes:

1. Daily follow-up
2. Weekly follow-up
3. Fortnightly follow-up
4. 4-weekly follow-up
5. Follow-up at the beginning of weeks 2 and 4, then every 4 weeks
6. Follow-up in the middle of weeks 2 and 4, then every 4 weeks
7. Follow-up at the end of weeks 2 and 4, then every 4 weeks

For each of these follow-up schemes, we fit the models to the data as described above. The different follow-up schemes affect the AIC such that is not possible to compare the fit of the models with different follow-up schemes based on the AIC as follow-up schemes with a longer time between visits result in a lower AIC (see **Table S24**).

**Comparison of the different follow-up schemes by AIC (Thailand-Myanmar data)**

| **Follow-up scheme** | **Model 1:** constant relapse rate | **Model 2:** temporal heterogeneity | **Model 3:** population heterogeneity |
|---|---|---|---|
| 1 | 8794 | 8492 | 8426 |
| 2 | 5865 | 5610 | 5544 |
| 3 | 4787 | 4596 | 4532 |
| 4 | 3712 | 3653 | 3606 |
| 5 | 4289 | 4040 | 3990 |
| 6 | 4602 | 4319 | 4264 |
| 7 | 5023 | 4721 | 4663 |

**Table S24** Comparison of the different follow-up schemes by AIC (Thailand-Myanmar data). The AICs are affected by the follow-up scheme such that the model cannot be compared using the AIC. We used different infection rates for the two studies for these model fits and 10 relapse risk groups for model 3.

The model fits for models 1 to 3 with the different follow-up schemes are shown in **Fig. S18**, **Fig. S19**, and **Fig. S20**, respectively. There is little qualitative difference between the model fits with the different follow-up schemes. For each of the seven proposed follow-up schemes, the conclusion that model 3 gives the best fit with the lowest AIC and that only models 2 and 3 show a biphasic decay in the survival curves with a steeper initial decay and then a slower decay holds. Hence, we choose the first follow-up scheme (daily follow-up) and use this follow-up scheme for all model comparisons and model fits.

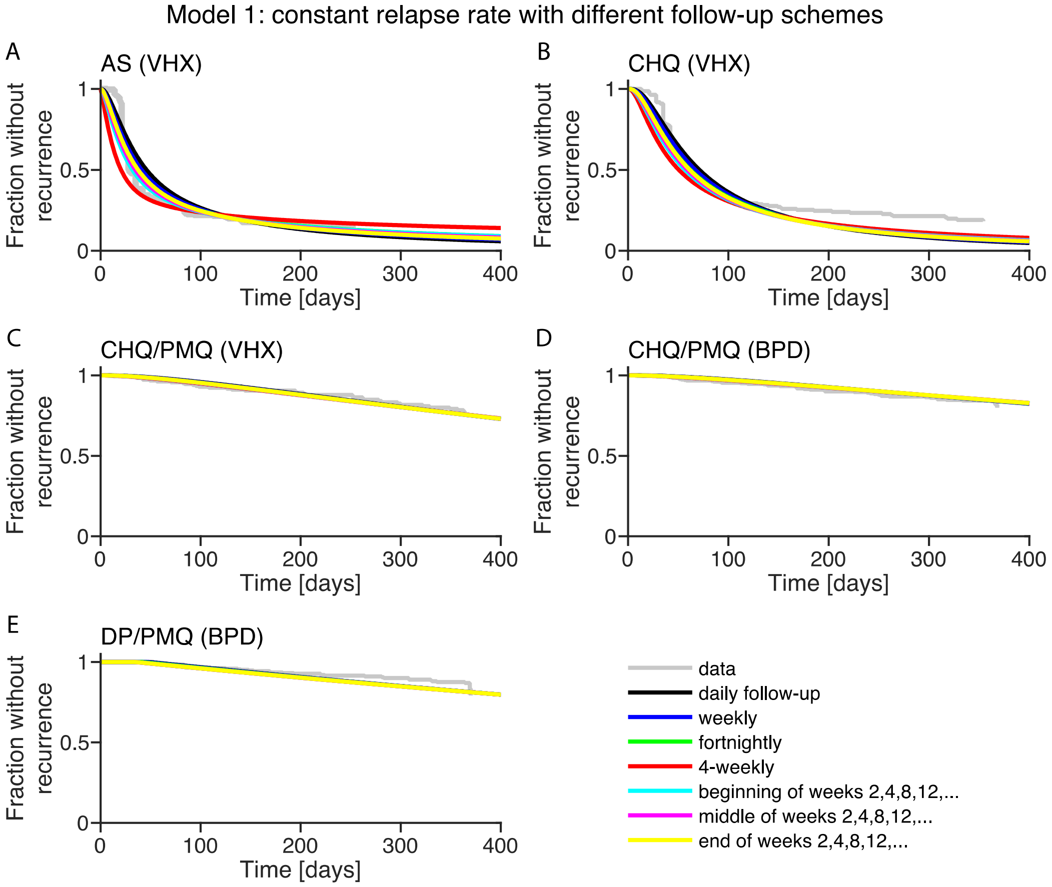


**Fig. S18** Fit of model 1 using different follow-up schemes for each antimalarial treatment and study in the Thailand-Myanmar data. The first recurrence data is shown in gray for comparison.

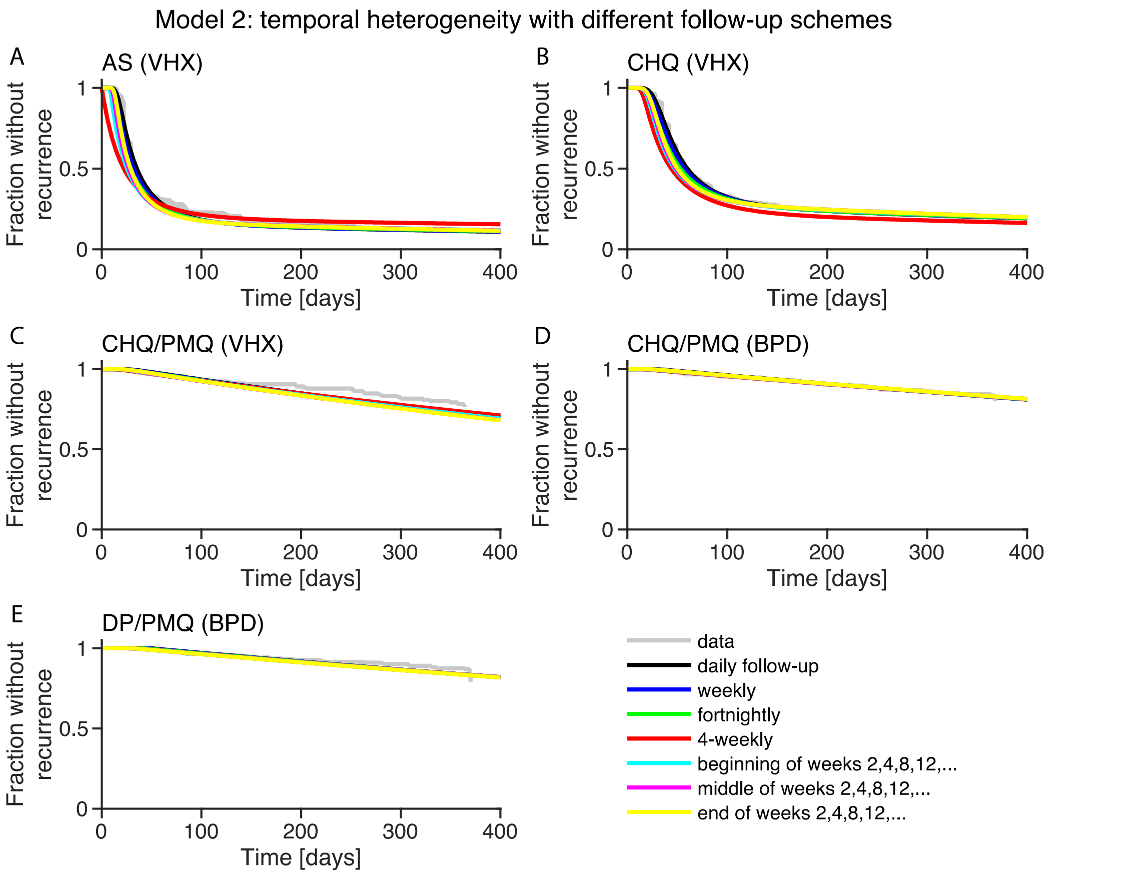


**Fig. S19** Fit of model 2 using different follow-up schemes for each antimalarial treatment and study in the Thailand-Myanmar data. The first recurrence data is shown in gray for comparison.

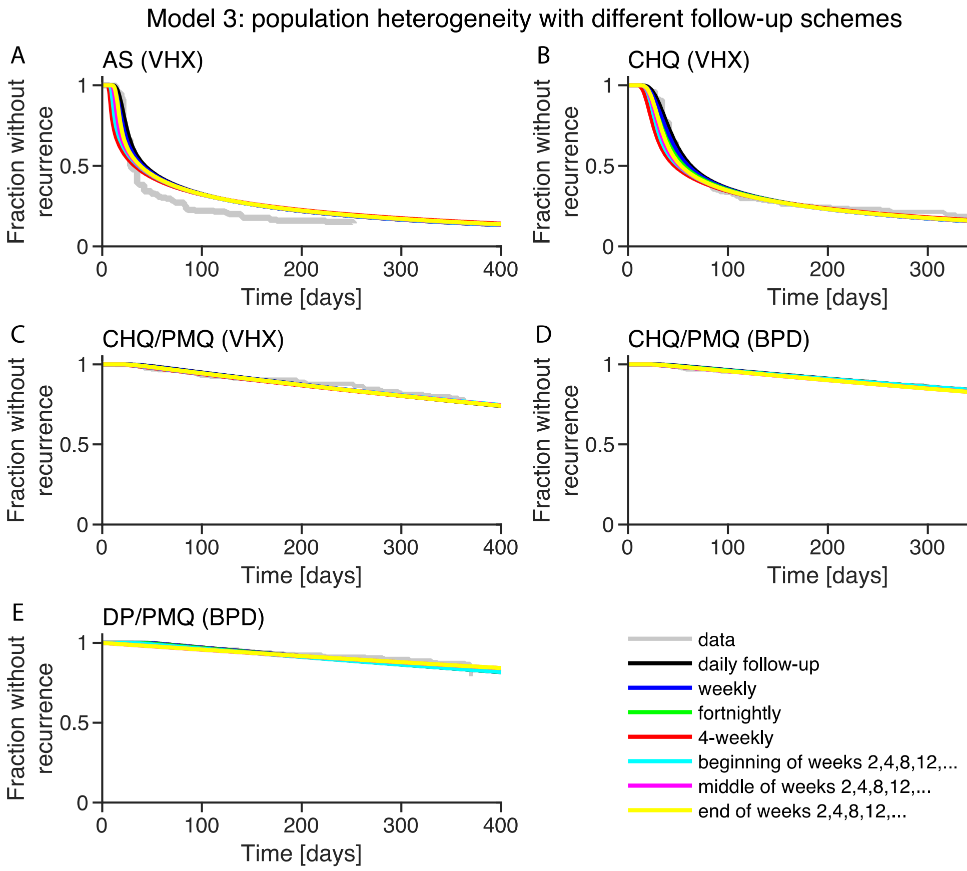


**Fig. S20** Fit of model 3 using different follow-up schemes for each antimalarial treatment and study in the Thailand-Myanmar data. The first recurrence data is shown in gray for comparison.

#### Different numbers of relapse risk groups in the population heterogeneity model for the Thailand-Myanmar data

In model 3, the relapse rate is constant but drawn from a lognormal distribution to model population heterogeneity. Individuals are grouped into $k$ relapse risk groups of equal size (see **Fig. S15**). We fit model 3 for $k = 5, 10, 15$, and 20 groups. Since the AICs are the same and the model fits are very similar (see **Table S25** and **Fig. S21**), we use $k = 10$ for all model fits, comparisons, and simulations.

**AICs for model 3 with different numbers of relapse risk groups (Thailand-Myanmar data)**

| Numbers of relapse risk groups | AIC |
|---|---|
| 5 | 8426 |
| 10 | 8426 |
| 15 | 8426 |
| 20 | 8426 |

**Table S25** Comparison of the AICs for model 3 with different numbers of relapse risk groups in the Thailand-Myanmar data. Model 3 was fit with two infection rates and daily follow-ups.

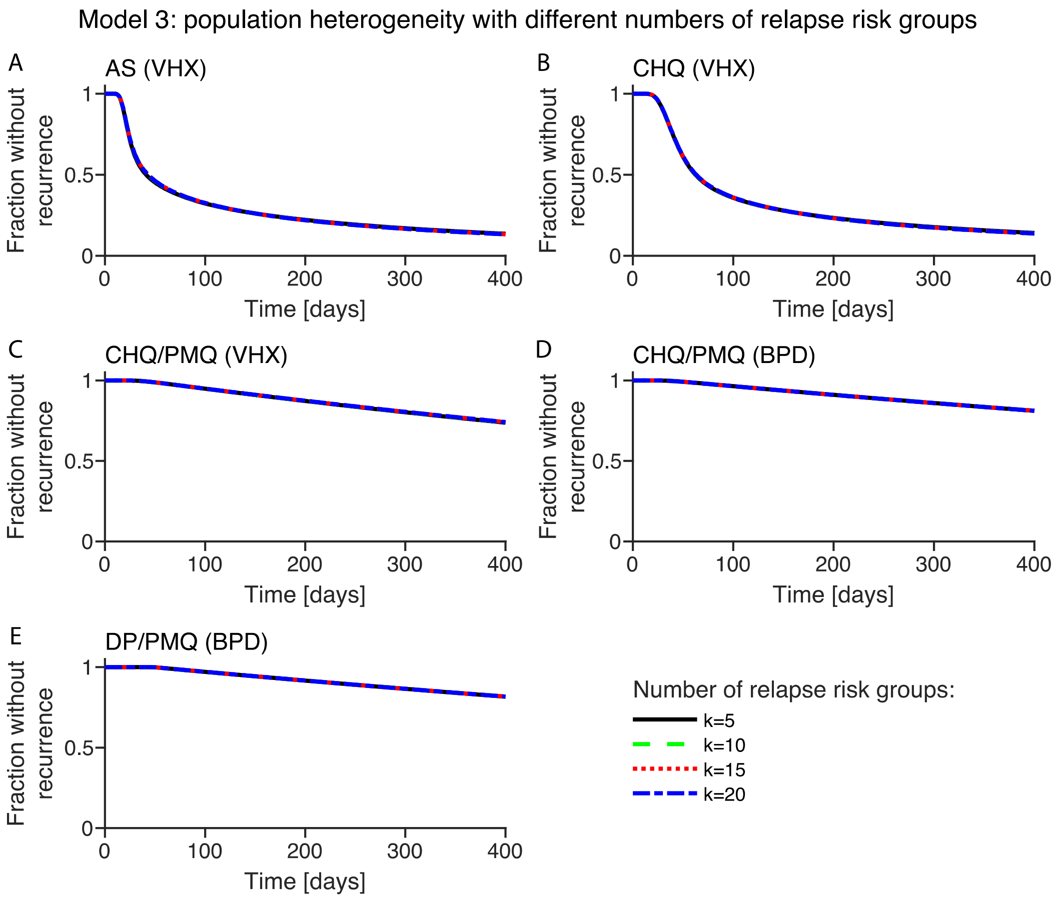


**Fig. S21** Comparison of the model fit of model 3 with different numbers of relapse risk groups (Thailand-Myanmar data). Each subfigure contains the best model fit of model 3 to one treatment group for different numbers of relapse risk groups, $k = 5, 10, 15$, and 20. For these model fits, we used different infection rates for the different studies and daily follow-up.

**Comparison of the population heterogeneity model fit to the Thailand-Myanmar data with lognormal, gamma, and exponential distribution for the relapse rates**

For the model comparisons, we used a lognormal distribution for relapse rates in the population heterogeneity model. We also fitted the population heterogeneity model using the gamma and exponential distributions as the distribution of relapse rates. The lognormal and the gamma distributions of relapse rates give a similar fit to the Thailand-Myanmar data, with the lognormal distributed relapse rates model having a slightly lower AIC than the model with gamma distributed relapse rates (see **Fig. S22**). Furthermore, both models have the same number of parameters and similar parameter value estimates (see **Table S26**). The exponential distribution has one less parameter but does not give as good a fit as either the lognormal or the gamma distribution.

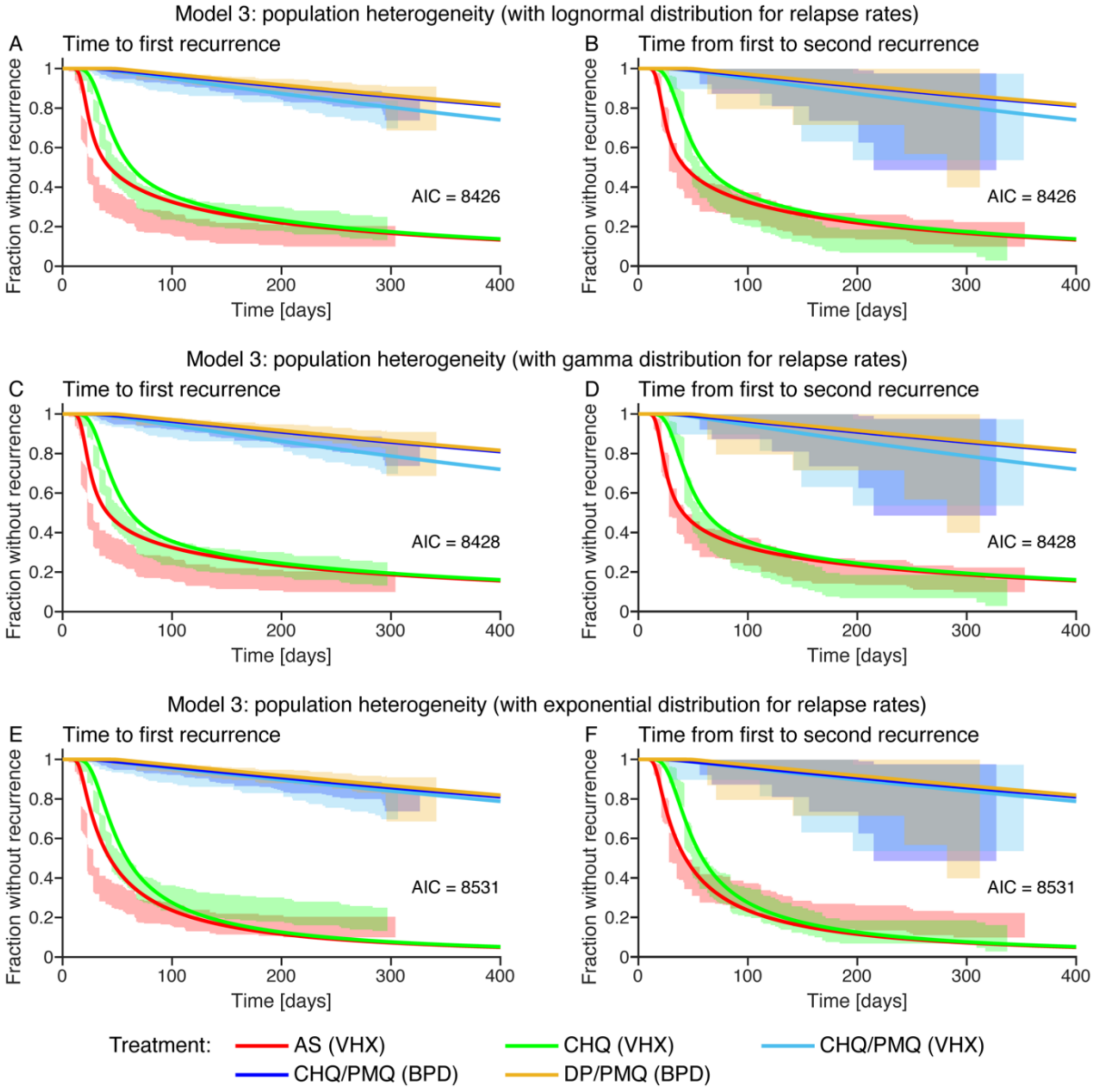


**Fig. S22** Fit of the population heterogeneity model with a lognormal, a gamma, and an exponential distribution of the relapse rates to the first and second recurrence time in the Thailand-Myanmar data. For these model fits, we used different infection rates for the different studies, daily follow-up, and 10 relapse risk groups.

**Parameters of model 3: population heterogeneity (Thailand-Myanmar data, lognormal, gamma, and exponential distribution of relapse rates)**

| Parameter | Lognormal d. | Gamma distr. | Exp. distr. |
|---|---|---|---|
| Mean of the logarithmic values of the drug washout time distribution for AS | 2.95 | 2.90 | 2.72 |
| Standard deviation of the logarithmic values of the drug washout time distribution for AS | 0.24 | 0.24 | 0.22 |
| Mean of the logarithmic values of the drug washout time distribution for CHQ | 3.53 | 3.51 | 3.34 |
| Standard deviation of the logarithmic values of the drug washout time distribution for CHQ | 0.33 | 0.34 | 0.31 |
| Mean of the logarithmic values of the drug washout time distribution for DP | 3.88 | 3.88 | 3.89 |
| Standard deviation of the logarithmic values of the drug washout time distribution for DP | 0.060 | 0.059 | 0.061 |
| Rate of new infections in the VHX study [per day] | 0.0008 | 0.0009 | 0.0006 |
| Rate of new infections in the BPD study [per day] | 0.0006 | 0.00058 | 0.0006 |
| Relapse rate distribution for AS and CHQ, parameter 1 | -3.73 | 0.30 | 0.035 |
| Relapse rate distribution for AS and CHQ, parameter 2 | 2.80 | 0.38 | - |

**Table S26** Parameter estimates for the parameters of the population heterogeneity model fit simultaneously to the first and second recurrence time in the Thailand-Myanmar data for a lognormal distribution of relapse rates and a gamma distribution of relapse rates. The parameter estimates are the maximum likelihood estimates for the model fits with different infection rates for the different studies, daily follow-up, and 10 relapse risk groups. Abbreviations: AS artesunate treatment, CHQ chloroquine treatment, DP dihydroartemisinin-piperaquine treatment, PMQ primaquine treatment, VHX Vivax History study, BPD Best Primaquine Dose study.

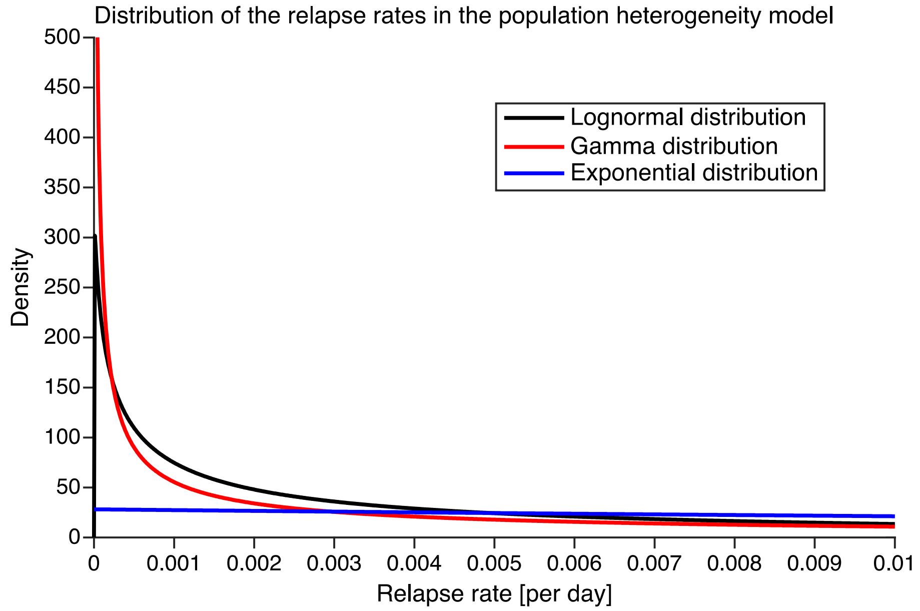


**Fig. S23** Comparison of the relapse rate distribution in the population heterogeneity model with a lognormal, a gamma, and an exponential distribution of relapse rates.
